# Sex-specific cortical brain differences in children at familial high risk for schizophrenia or bipolar disorder

**DOI:** 10.1101/2025.04.24.25326262

**Authors:** Kathrine Skak Madsen, William F.C. Baaré, Enedino Hernandez-Torres, Kit Melissa Larsen, Adam Kaminski, Line Korsgaard Johnsen, Nicoline Hemager, Maja Gregersen, Julie Marie Brandt, Mette Falkenberg Krantz, Nanna Weye, Anne Søndergaard, Aja Neergaard Greve, Christina Bruun Knudsen, Anna Krogh Andreassen, Lotte Veddum, Torben E. Lund, Ole Mors, Anne Amalie Elgaard Thorup, Leif Østergaard, Merete Nordentoft, Hartwig R. Siebner

## Abstract

**Background:** Schizophrenia (SZ) and bipolar disorder (BP) are severe psychiatric disorders with neurodevelopmental underpinnings. Familial high risk (FHR) is the strongest predictor of developing SZ and BP. Children at FHR offer a unique opportunity to identify early brain markers of vulnerability. However, previous studies often span wide age ranges and neglect sex differences, despite evidence of distinct sex-specific brain developmental trajectories. We investigated sex-specific group differences in brain morphometry among 11-12-year-old children at FHR for SZ (FHR-SZ) or BP (FHR-BP).

**Methods:** This study included 278 children from the Danish High Risk and Resilience Study (VIA11): 101 FHR-SZ, 64 FHR-BP, and 113 population-based controls (PBCs). Groups matched on age and sex. Structural MRI scans were acquired on 3T scanners at two sites. Brain volume, cortical volume, surface area, and cortical thickness were extracted using FreeSurfer.

**Results:** Significant group-by-sex interactions were observed for brain volume, cortical volume, and surface area (eta^2^=0.030-0.038; p=0.006-0.016). Males at FHR-SZ exhibited smaller brain volume, cortical volume, and surface area than PBC males (Cohen’s *d*=-0.624--0.489; p=0.002-0.015), while FHR-BP females had larger brain and cortical volumes than PBC females (Cohen’s *d*=0.525-0.537; p=0.017-0.020). No significant differences were observed for cortical thickness (p>0.210).

**Conclusions:** Children at FHR-SZ and FHR-BP exhibited sex-specific morphometric differences, potentially reflecting sex-specific endophenotypic markers of risk. Our findings highlight the importance of incorporating sex as a factor in neurodevelopmental psychiatric research. Longitudinal studies are needed to track how these neuroanatomical differences evolve over time and to evaluate their predictive value for transition to SZ or BP.

## Introduction

Schizophrenia (SZ) and bipolar disorder (BP) are debilitating psychiatric disorders that may cause profound personal suffering and emotional distress for those affected and their families, and substantial societal costs (1). A family history of severe mental illness is the strongest risk factor for developing severe mental disorders (2,3). More than 50% of children born to parents with SZ or BP will develop any mental illness (4,5), and 15-40% of children at familial high risk (FHR) for schizophrenia (FHR-SZ) or bipolar disorder (FHR-BP) are estimated to develop a psychotic or bipolar disorder in adulthood (3,6). Identifying vulnerable individuals before the onset of illness is critical for early prevention. Enriched cohorts, such as children at FHR, offer a unique opportunity to investigate the neurodevelopmental processes that precede severe mental disorders.

SZ and BP are hypothesized to be neurodevelopmental disorders associated with atypical brain development and alterations in brain structure and function (7–10), which may contribute to profound cognitive, emotional, and behavioral challenges. Individuals with SZ or BP, and, to a lesser degree, their first-degree relatives, consistently exhibit widespread brain morphological alterations (11–14). SZ is associated with reduced grey and white matter volumes, increased ventricular size, and pronounced cortical thinning (14–17), with similar but more subtle alterations observed in first-degree relatives (13,18). Individuals with BP exhibit more modest reductions in brain and cortical volumes and cortical thickness (11,13,15,16,19), while their first-degree relatives show larger intracranial, brain, and cortical volumes and surface area (13,20,21).

Studies of children and adolescents at FHR-SZ or FHR-BP have identified brain morphological differences that may serve as early biomarkers of neurodevelopmental risk (22–26). Children and adolescents at FHR-SZ (aged 8-23) exhibit smaller brain and gray matter volumes, thinner cortices, and reduced surface area compared to FHR-BP adolescents and controls (22–24), with differences in surface area and gray matter volume linked to subsequent psychosis onset(25). FHR-BP children and adolescents (aged 6-23) show subtler brain alterations (24–26), including larger ventricular size (22). Although valuable, previous studies often examine small sample sizes with broad age ranges (6-23 years), spanning up to a decade within a single cohort, thus including individuals at different stages of brain maturation. Additionally, most prior FHR studies in children and adolescents did not examine sex differences, despite known sex differences in brain maturational trajectories in this age range (27–30). Hormonal influences have been hypothesized to contribute to divergent neurodevelopmental trajectories associated with SZ and BP, underscoring the need for research on sex differences (31). These constraints hinder the ability to pinpoint age- and sex-specific neurodevelopmental processes crucial for the later development of SZ or BP. Consequently, the timing and nature of the brain structural changes remain poorly understood, particularly before the emergence of SZ or BP symptoms.

This study aims to address these gaps by utilizing the baseline neuroimaging data from the prospective *Danish High Risk and Resilience Study (VIA)* (32), which follows a large cohort of children born to parents with SZ, BP, or neither of these disorders. We investigated whether 11-to-12-year-old children at FHR-SZ or FHR-BP differed from population-based controls (PBC) in brain volume, cortical volume, cortical thickness, and surface area. Given the established sex differences in brain maturational trajectories (27–29) as well as in clinical presentation and symptoms (31,33,34), we hypothesized that group differences would be sex-specific. Therefore, we first specifically tested for group-by-sex differences, followed by an examination of overall group differences. Finally, we conducted exploratory parcel-based analyses to identify cortical regions with significant differences in brain morphometry.

## Methods

### Participants

A total of 522 7-year-old children were enrolled in the *Danish High-Risk and Resilience study (VIA 7)* (32,35), a prospective, longitudinal, register-based cohort study of children born to at least one parent with SZ (FHR-SZ, n = 202) or BP (FHR-BP, n = 120) or neither of these disorders (PBC, n = 200). SZ was defined as ICD-10 codes F20, F22, and F25 or ICD-8 codes 295, 297, 298.29, 298.39, 298.89, and 298.99. Bipolar disorder was defined as ICD-10 codes F30 and F31 or ICD-8 codes 296.19 and 296.39. Children were recruited from the Danish Psychiatric Central Research Register (36) and the Danish Civil Registration System. Permission to retrieve the cohort from the Danish national registers was granted by the Danish Ministry of Health. At enrollment, PBC children were matched to the FHR-SZ children in age, sex, and municipality (37,38). A flowchart detailing the VIA recruitment process, from data extraction from the Danish registries to enrolment in the VIA study, has been presented elsewhere (39).

A total of 465 children (FHR-SZ: n = 179, FHR-BP: n = 105, PBC: n = 181) participated in the VIA 11 study when they were 11-12 years old. The present study focuses on the structural magnetic resonance imaging (MRI) scans acquired in the VIA 11 study and included 278 children (FHR-SZ: n = 101; FHR-BP: n = 64, PBC: n = 113). An overview of the inclusion/exclusion procedure is shown in the flowchart in Figure S1. Further study details are in the VIA study protocols (32,35,40). Prior to participation, we obtained written informed consent from the children’s legal guardian(s) and oral assent from the children. The VIA 11 study was approved by the National Committee on Health Research Ethics (Protocol: H-16043682) and the Danish Data Protection Agency (ID: RHP-2017-003, I-suite: 05333) and conducted in accordance with the Declaration of Helsinki.

### Clinical variables

Children’s level of behavioral problems was assessed using the Child Behavior Checklist (CBCL) school-age version (41) filled in by the caregiver. The Children’s Global Assessment Scale (CGAS (42)) evaluated the children’s level of general functioning in the preceding month based on caregiver-child interviews. Lifetime axis-I disorders (excluding elimination disorders, transient/unspecified tics, and specific phobias) were assessed using the semi-structured interview for Affective Disorders and Schizophrenia for School-Age Children - Present and Lifetime Version (K-SADS-PL (43)) followed by a clinical conference with a specialist in child and adolescent psychiatry (AT)^6^. There were no diagnoses of mania (DSM 296). Two children met the criteria for an axis-I psychotic disorder (DSM 298.9; none with 298.8/297.1/292.30/295.90) (5). A Personal and Social Performance (PSP) Scale interview assessed the primary caregiver’s level of functioning (44).

### MRI protocol

Participants underwent harmonized anatomical and functional whole-brain MRI for approximately 75 minutes at the Danish Research Centre for Magnetic Resonance (DRCMR) in Copenhagen or the Centre of Functionally Integrative Neuroscience (CFIN) in Aarhus. Participants were scanned on a 3T Siemens Magnetom Prisma Fit MR scanner (Siemens, Erlangen, Germany) using a 64-channel head coil at DRCMR and on a Siemens 3T Magnetom Skyra with a 32-channel head coil at CFIN. T1-weighted images were derived from 3D magnetization prepared 2 rapid acquisition gradient echoes (MP2RAGE) sequences with fat image navigators for retrospective motion correction (45) using similar parameters, except for minor differences in TE and acquisition time (TR = 6500 ms, TI_1_ = 700 ms, TI_2_ = 2800 ms, TE_DRCMR/CFIN_ = 3.49/3.46 ms, flip angle = 4°/6°, FoV = 260 × 260 mm, 192 sagittal slices, voxel size = 0.9 x 0.9 x 0.9 mm^3^, acquisition time_DRCMR/CFIN_ = 12:39/12:28). Prior to analysis and blind to group status, images were visually inspected to ascertain image quality and excluded if of inadequate quality (Figure S1). Indications of clinical findings were consulted with a neuroradiologist and referred to medical auspices, if relevant.

### FreeSurfer preprocessing, extraction of brain measures, and quality assessment

T1-weighted images were processed using FreeSurfer (46–48) version 7.1.1. Cortical grey matter parcellations were based on surface-based nonlinear registration to the Desikan-Killiany atlas, yielding estimates for 34 parcels in each hemisphere (49). We extracted total brain volume and measures of cortical gray matter volume, cortical thickness, and white matter surface area from the whole cerebral cortex and each parcel. The samseg (50) processing stream in FreeSurfer was used to derive a robust measure of intracranial volume (ICV).

FreeSurfer outputs were visually inspected for quality using an in-house graphical user interface. FreeSurfer outputs of questionable quality were discarded. Finally, we extracted the Euler number for each participant and used it as a quantitative measure of image quality (51) in our statistical analyses. Three scans were excluded due to poor quality, determined by a Euler number more than three standard deviations below the mean, after adjusting for age, sex, and site.

### Statistical analyses

Analyses were conducted in R (version 4.0.4), primarily using the packages *car*, *lsmeans*, and *BayesFactor*. Group differences in demographic and clinical variables were tested using one-way analysis of variance (ANOVA) or chi-square tests, followed by pairwise comparisons. Drop-out analyses employed t-tests and chi-square tests. To test our hypotheses of sex-specific group differences in brain morphometry, we performed analysis of covariance (ANCOVA) to examine group-by-sex interactions in brain volume, cortical volume, surface area, and mean cortical thickness, controlling for age, sex, site, and Euler number. If the group-by-sex interaction was not significant, it was removed, and the main effect of group was assessed independently. We used Benjamini-Hochberg false discovery rate (FDR) to correct for multiple comparisons across the four brain measures, using an FDR-corrected threshold of q = 0.05. In addition to reporting p-values, we supplemented our primary statistical models with a Bayesian approach, presenting Bayes factors (BF_10_) to quantify the level of support for the alternative hypothesis (H_1_) compared to the null hypothesis (H_0_). Following the standard interpretation, BF_10_ ≤ 1 indicates no evidence, 1-3 indicates anecdotal evidence, 3-10 indicates moderate evidence, 10-30 indicates strong evidence, 30-100 indicates very strong evidence, and BF_10_ > 100 indicates decisive evidence (52,53).

Significant results were followed up with additional analyses to further elucidate the observed group-by-sex associations, using an uncorrected p-value of 0.05. First, we examined whether observed group-by-sex effects persisted when controlling for ICV or height. Subsequent analyses were conducted separately in females and males. We conducted pairwise group comparisons using t-tests on the estimated marginal means from *lsmeans*. Post hoc, we examined whether group differences in brain measures were related to lifetime axis-I diagnosis by i) controlling for diagnosis and ii) stratifying FHR-SZ, FHR-BP, and PBC groups into subgroups *with* (+) or *without* (-) an axis-I diagnosis.

Finally, parcel-based analyses explored how group differences in cortical volume, surface area, and cortical thickness were distributed across the cortex in males and females. We conducted pairwise t-tests on lsmeans for the 68 Desikan parcels, applying FDR correction for 68 analyses.

## Results

### Demographic and clinical characteristics

Demographic and clinical characteristics and group differences are presented in Table 1. Groups did not differ significantly in sex, age, or scan site. FHR groups had significantly higher CBCL scores and lower CGAS scores than PBCs, indicating more behavioral problems and lower general functioning. They also had a higher prevalence of any lifetime axis-I diagnosis, as previously reported for the whole VIA11 cohort (54). Drop-out analysis revealed that included FHR-SZ participants had higher CGAS and lower total CBCL scores, and their primary caregiver had higher PSP scores than those not included (p < 0.028), i.e., included children and their primary caregiver were better functioning. No other drop-out differences were observed (Table S1-S2).

**Table 1.** Demographic and clinical characteristics and group differences between the FHR-SZ, FHR-BP, and population-based controls (PBC) groups.

|  | FHR-SZ | FHR-BP | PBC | p | Pairwise Comparisons |  |  |
| --- | --- | --- | --- | --- | --- | --- | --- |
|  |  |  |  |  | p |  |  |
|  |  |  |  |  | FHR-SZ<br>vs.<br>PBC | FHR-BP<br>vs.<br>PBC | FHR-SZ<br>vs.<br>FHR-BP |
| Children, N | 101 | 64 | 113 |  |  |  |  |
| Females, N (%) | 50 (49.5) | 32 (50.0) | 56 (49.6) | 0.998 | - | - | - |
| Age, mean (SD) | 12.11 (0.28) | 12.07 (0.31) | 12.13 (0.27) | 0.436 | - | - | - |
| Site, DRCMR, N (%) | 52 (51.5) | 30 (46.9) | 45 (39.8) | 0.226 | - | - | - |
| CBCL Total, mean (SD)<sup>a</sup> | 20.88 (17.89) | 20.88 (21.24) | 11.65 (10.68) | **<0.001** | **<0.001** | **0.002** | 0.999 |
| CGAS, mean (SD)<sup>a</sup> | 67.84 (14.70) | 69.45 (15.44) | 75.86 (14.22) | **<0.001** | **<0.001** | **0.007** | 0.507 |
| Lifetime axis-I disorder, N (%)<sup>a</sup> | 51 (51.5) | 31 (48.4) | 31 (27.9) | **0.001** | **<0.001** | **0.010** | 0.823 |

Group differences in age, CBCL, and CGAS were tested using a one-way ANOVA. Group differences in sex, site, and prevalence of lifetime axis-I disorders (excluding elimination disorders, transient/unspecified tics, and specific phobias) were tested using a chi-square test. Significant p-values are shown in bold. Lower CBCL scores indicate more behavioral problems, while higher CGAS scores indicate better global functioning. Abbreviations: FHR-SZ: Familial high risk of schizophrenia, FHR-BP: Familial high risk of bipolar disorder, PBC: Population-based controls, CBCL: Child Behavior Check List, CGAS: Children's Global Assessment Scale. <sup>a</sup> Missing data: CBCL (n=6), CGAS (n=2), axis-I disorders (n=4).

### Group-by-sex interaction and group differences on global brain measures

As hypothesized, we observed significant group-by-sex interaction effects on brain volume (eta^2^ = 0.038, F = 5.270, p = 0.006, q = 0.021, BF_10_ = 8.362), cortical volume (eta^2^ = 0.031, F = 4.364, p = 0.014, q = 0.021, BF_10_ = 3.523), and surface area (eta^2^ = 0.030, F = 4.220, p = 0.016, q = 0.021, BF_10_ = 2.982), with no main effects of group (Table S3), suggesting sex-specific group differences in the brain measures. For mean cortical thickness, no significant group-by-sex (eta^2^ = 0.002, F = 0.20, p = 0.801, BF_10_ = 0.088) or group (eta^2^ = 0.019, F = 2.775, p = 0.064, BF_10_ = 0.507) effects were observed, with Bayes factors indicating strong to anecdotal evidence against differences (Tables S3-S4).

Adjusting for height did not alter the group-by-sex effects in brain volume, cortical volume, and surface area (F > 3.898, p < 0.021), suggesting that these were not linked to differences in overall growth (Table S5). None of the group-by-sex effects survived adjustment for ICV (Table S6), likely reflecting the interdependence of brain growth and skull growth during childhood (55,56) (Supplementary Results).

To facilitate comparison with previous studies, we conducted post hoc analyses on the main effect of group, excluding the group-by-sex interaction (Table S4). We found significant group differences in brain volume (eta^2^ = 0.031, F = 4.288, p = 0.015, BF_10_ = 1.885) and cortical volume (eta^2^ = 0.028, F = 3.937, p = 0.021, BF_10_ = 1.498), though Bayes factor provided only anecdotal evidence. We found no significant group differences for surface area or cortical thickness (p > 0.064), and no evidence from the Bayes factor (BF_10_ < 0.507, Table S4). Overall, the results suggest that sex-specific group differences underlie the observed findings.

### Group differences in the global brain measures in females and males

To disentangle the observed group-by-sex effects, we conducted pairwise group analyses in females and males separately (Figure 1, Table S7). Females at FHR-BP exhibited significantly larger brain and cortical volumes than PBC females (p < 0.020), but Bayesian evidence was anecdotal (BF_10_ < 2.062). Males at FHR-SZ had significantly smaller brain and cortical volumes than PBC and FHR-BP males (p < 0.013) with moderate to strong Bayesian evidence (BF_10_ range = 5.993-20.623). Males at FHR-SZ also displayed reduced surface area compared with PBC males (p = 0.015), though Bayesian evidence was anecdotal (BF_10_ = 2.473). No other significant group differences were observed.

**Figure 1.**
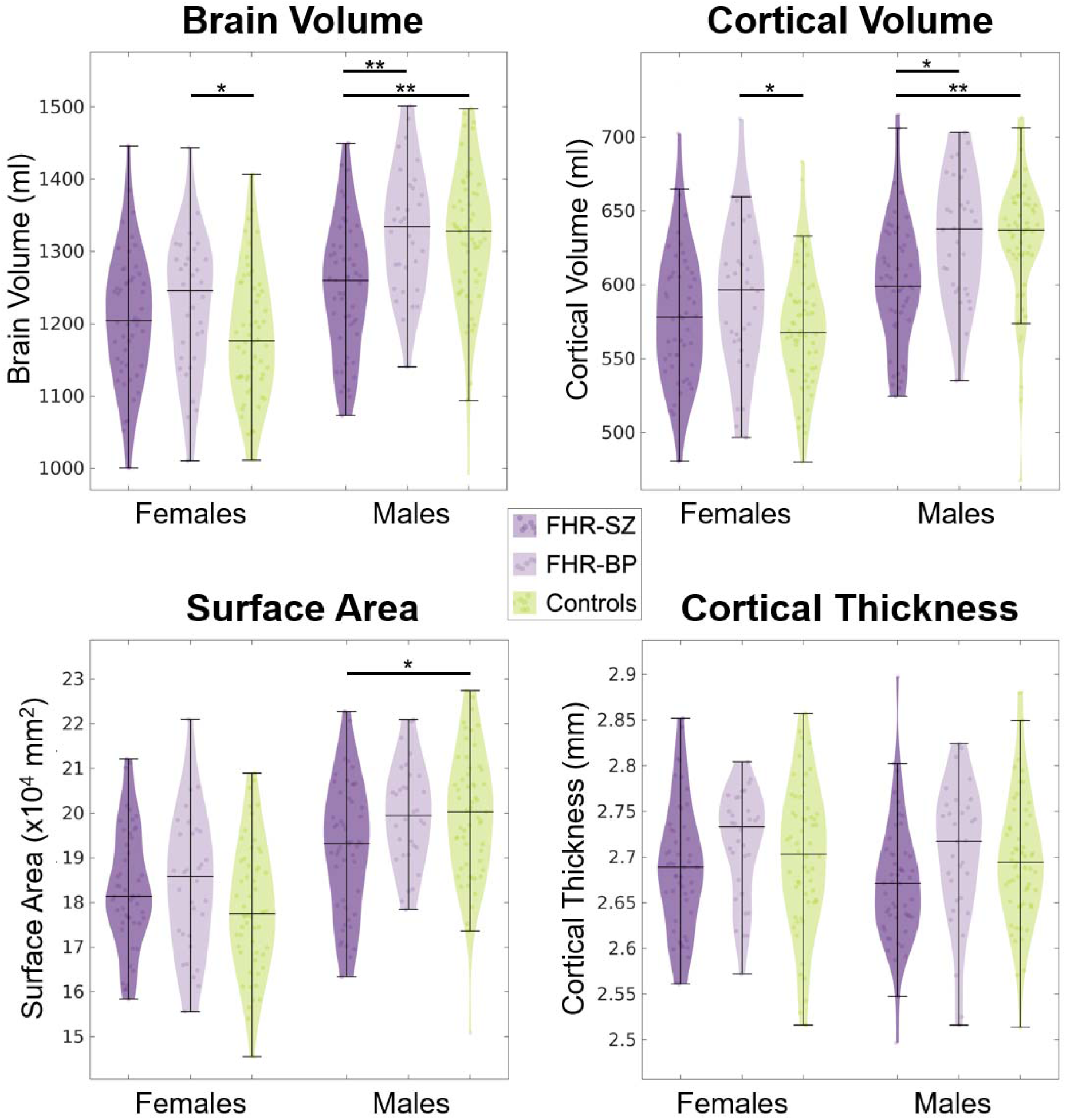
Violin plots displaying brain volume, cortical volume, surface area, and mean cortical thickness for females and males. The familial high risk for schizophrenia (FHR-SZ) or bipolar disorder (FHR-BP) groups are shown in, respectively, dark and light purple, while the population-based controls are shown in green. The horizontal lines indicate the median and the data range (excluding outliers). The dots within the violin plots show the individual data points. Number of male/female children in each group: FHR-SZ: n=51/50, FHR-BP: n=32/32, PBC: n=57/56. Statistical pairwise group differences are indicated at the 0.05 (*) and 0.01 (**) uncorrected significance levels.

### Group differences in global brain measures: role of lifetime axis-I diagnosis

To assess whether group differences were associated with axis-I diagnosis, we included it as a covariate. FHR-SZ males still displayed smaller brain and cortical volumes than FHR-BP and PBC males (p < 0.028), while surface area was no longer significant (p = 0.073), suggesting the volume reductions were not mediated by axis-I diagnosis (Table S8). In females, the previously observed larger brain and cortical volumes in FHR-BP versus PBC females were no longer significant after accounting for axis-I diagnosis (p > 0.058, Table S8)

Next, we stratified the groups by lifetime axis-I diagnosis status to explore differences between children *with* and *without* axis-I diagnosis (Figure 2, Table S9). We did not observe significant differences between males *with* or *without* an axis-I diagnosis (p > 0.354), suggesting axis-I diagnosis was not associated with volume or surface area variation. In females, brain and cortical volumes were marginally larger in FHR-SZ+ and FHR-BP+ compared to FHR-SZ-and FHR-BP-, with a significant difference only between the FHR-SZ subgroups for brain volume (p = 0.049).

**Figure 2.**
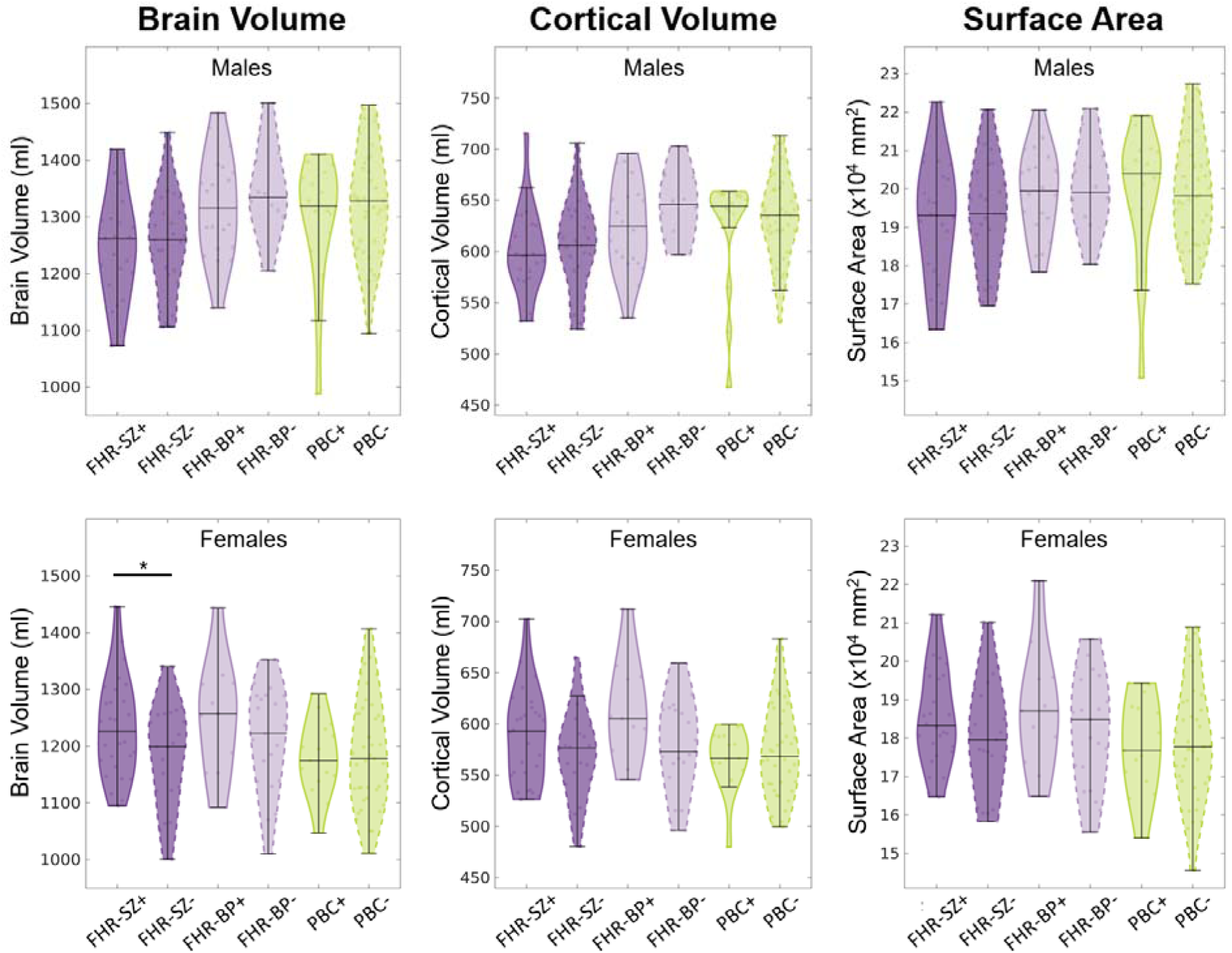
Violin plots displaying brain volume, cortical volume, and surface area for males (top) and females (bottom), with the groups stratified by children *with* (+) and *without* (-) a lifetime axis-I diagnosis. The familial high risk for schizophrenia (FHR-SZ) or bipolar disorder (FHR-BP) groups are shown in, respectively, dark and light purple, and the population-based controls are shown in green. The horizontal lines indicate the median and the data range (without outliers). The dots within the violin plots show the individual data points. Number of male/female children in each group: FHR-SZ+: n=23/28, FHR-SZ-: n=27/21, FHR-BP+: n=20/11, FHR-BP-: n=12/21, PBC+: n=16/15, PBC-: n=39/41.Statistical pairwise group differences are indicated at the 0.05 (*) uncorrected significance levels.

### Group differences in cortical measures across the cortex in females and males

We investigated how group differences in cortical volume and surface area were distributed across the cortex. Effect sizes are visualized on brain maps (Figure 3) and bar plots (Figures S3-S4). Exploratory analyses of cortical thickness are presented in Figures S2 and S5.

**Figure 3.**
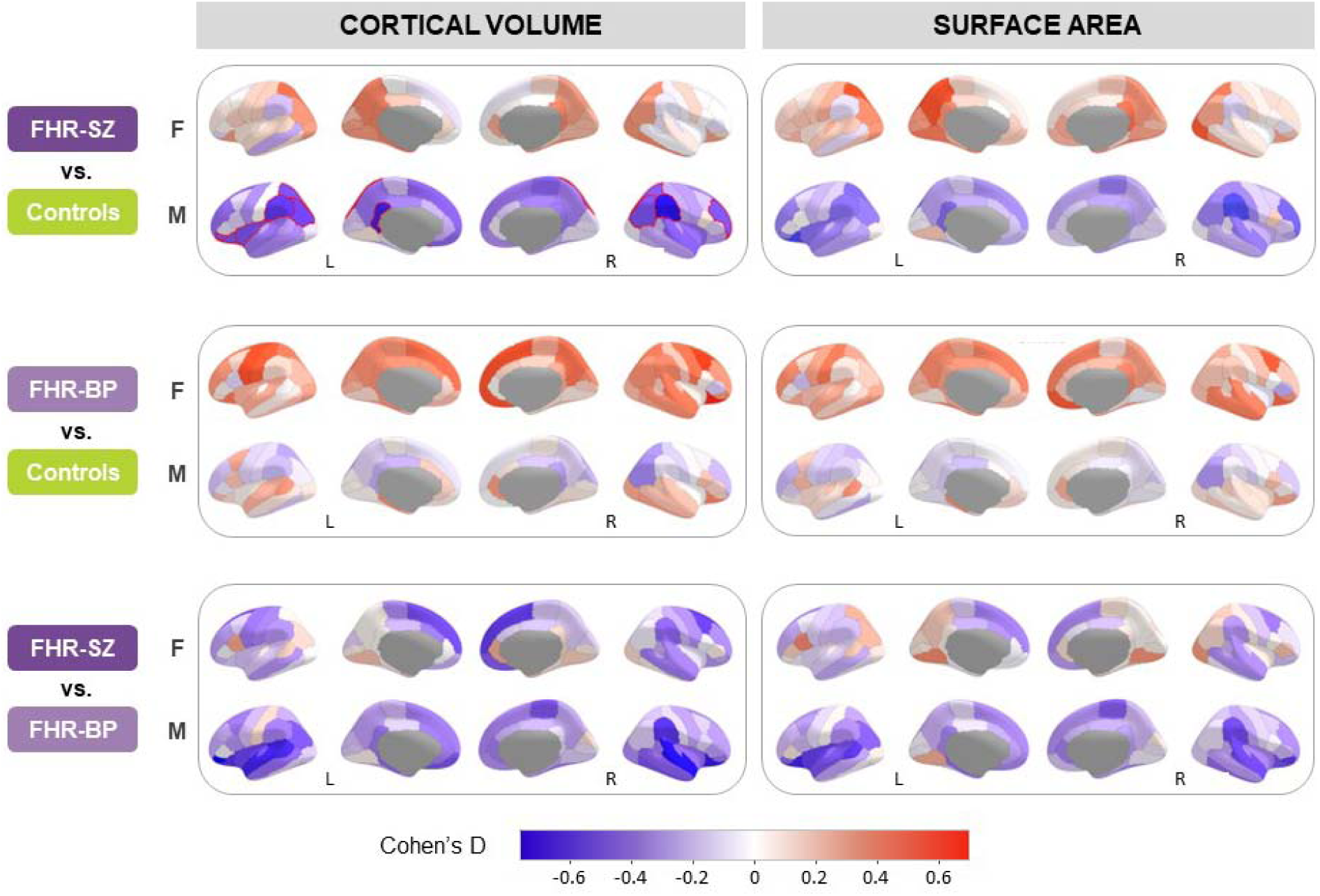
Effect size maps displaying the group difference in cortical volume (left) or surface area (right) between children at familial high risk for schizophrenia (FHR-SZ, top) or bipolar disorder (FHR-BP, middle) vs. population-based controls and between FHR-SZ vs. FHR-BP (bottom) for females (F) and males (M). Red shades indicate regions where FHR groups or FHR-SZ have larger cortical volume or surface area than, respectively, controls or FHR-BP. Blue shades indicate regions where FHR groups or FHR-SZ have smaller cortical volume or surface area than controls or FHR-BP. Red lines bordering the parcels indicate parcels that are significant after FDR correction (seen for cortical volume in the comparison between FHR-SZ vs. PBC males).

Males at FHR-SZ showed widespread smaller cortical volume and surface area than PBC males. The largest effect sizes for cortical volume were in the bilateral supramarginal, inferior, and superior parietal cortices, left insula, pars orbitalis, lateral orbitofrontal, and isthmus cingulate, and right rostral middle frontal cortices (Figure 3, Figures S3-S4). These regions remained significant after FDR correction. In contrast, FHR-SZ females generally showed small effect sizes relative to PBC females, with medium-sized increases (Cohen’s *d* > 0.5) in surface area observed in left precuneus, left superior parietal, right temporal pole, and right lateral occipital cortices.

FHR-BP females exhibited widespread increases in cortical volume and surface area relative to PBC females, especially in bilateral superior frontal, right orbitofrontal, right caudal middle frontal, and left pre- and postcentral cortices (Figure 3, Figures S3-S4), though none survived FDR correction. FHR-BP males showed no significant differences compared to PBC males.

Relative to FHR-BP, the FHR-SZ group exhibited widespread smaller cortical volume and surface area, with stronger effect sizes in males than females. In males, the largest effect sizes were in the bilateral lateral temporal, pars orbitalis, and orbitofrontal regions, left insula, left interior parietal, right supramarginal, and right paracentral cortices. In females, the largest effect sizes were in the frontal cortices, including bilateral superior, right caudal middle, right medial orbitofrontal, and right frontal pole. None of these differences survived FDR correction.

## Discussion

In the largest MRI study to date investigating 11-to-12-year-old children at FHR-SZ and FHR-BP, we identified novel sex-specific global volumetric differences. Our findings expand prior research by suggesting sex-specific brain differences in children at FHR-SZ or FHR-BP, an aspect that most previous FHR studies have not examined. Specifically, males at FHR-SZ exhibited smaller brain and cortical volumes and reduced surface area compared to male PBCs, while females at FHR-BP showed larger brain and cortical volumes than female PBCs. These differences were widespread across the cortex. Notably, we observed no evidence for differences in cortical thickness, as reported in adults with SZ and BP (11,12,14).

The reduced brain and cortical volumes in males at FHR-SZ align with studies reporting similar findings in 8-to-23-year-olds at FHR-SZ (22) and first-degree relatives (aged 6-70 years) of individuals with SZ (13), regardless of sex. These differences were not explained by lifetime axis-I disorders, suggesting they may reflect endophenotypic markers of familial risk. Additionally, sex-specific volumetric brain differences (in males but not females) have been observed in neonates at FHR-SZ (57), though the direction varies with age. Emerging evidence suggests that genetic factors influencing brain morphology in SZ may show sex-specific effects, potentially contributing to our findings (58,59). We observed the most pronounced reductions in cortical volume and surface area in the parietal, insula, and frontal cortices, aligning with recent findings of reduced frontal and parietal surface area in 10-to-20-year-old adolescents at FHR-SZ (23). Together, these findings point to sex-specific neurodevelopmental differences preceding the potential onset of severe mental illness, suggesting cortical measures as potential markers of vulnerability. Longitudinal studies are needed to elucidate neurodevelopmental pathways, assess the clinical implications, and identify individuals at the highest risk for transitioning to severe mental illness.

Females at FHR-BP exhibited larger brain and cortical volumes than female PBCs, consistent with studies showing larger ICV and brain volumes in first-degree relatives of individuals with BP, regardless of sex (13,21). The increased volume may represent a structural marker of genetic vulnerability to BP. This is supported by a twin study including both sexes, showing a positive correlation between genetic risk for BP and ICV (60), along with brain growth and intracranial growth being closely interlinked during childhood (55,56,61). Surprisingly, FHR-BP females with an axis-I disorder showed marginally larger cortical volumes than those without, aligning with recent research reporting larger regional cortical volumes in 10-to-20-year-old adolescents at FHR-BP, particularly those exhibiting subclinical manic and depressive symptoms (62). The mechanisms underlying larger brain volumes in FHR-BP females warrant further exploration. Given the small FHR-BP sample size and anecdotal Bayes evidence, further studies in larger longitudinal cohorts are needed to validate these findings and assess their potential as early markers of vulnerability.

### Strengths and limitations

This study represents the largest MRI investigation to date of same-aged children at FHR-SZ or FHR-BP, recruited through national registries. Providing a robust dataset with greater statistical power, our study offers novel insights into the interplay of familial risk, neuroanatomy, and sex. Examining a narrow age range around puberty pinpoints neuroanatomical differences during this sensitive developmental period, enabling identification of brain differences prior to the onset of severe mental illness. The smaller FHR-BP sample size may limit the statistical power to detect subtle differences and increase the risk of spurious results, warranting cautious interpretation and replication of the findings. Slightly higher attrition among the most vulnerable children at FHR-SZ may introduce selection bias, though it is unlikely to explain the findings and may even have underestimated the group effects. By using a binary classification of axis-I without accounting for clinical heterogeneity within and across groups, we cannot identify nor control for brain morphometric patterns linked to specific psychopathological profiles. Finally, the study’s cross-sectional nature precludes causal inferences and tracking of neurodevelopmental trajectories.

## Conclusions

This study identified sex-specific cortical differences in 11-to-12-year-old children at FHR-SZ or FHR-BP, pinpointing potential early neurodevelopmental markers of familial risk for severe mental disorders. Males at FHR-SZ exhibited smaller brain and cortical volumes and surface area, independent of lifetime axis-I disorder, potentially reflecting endophenotypic markers of biological vulnerability prior to the onset of SZ or BP. In contrast, females at FHR-BP showed larger brain and cortical volumes, warranting further investigation due to the smaller sample. Our findings underscore the importance of investigating sex differences in developmental neuropsychiatric research. Longitudinal studies are essential to track how these neuroanatomical differences evolve over time and to assess their clinical implications and predictive value for SZ and BP.

## Data Availability

Due to Danish legislation, we cannot offer access to data unless there is a specific collaboration agreement with the research group.

## Acknowledgements

We are grateful to the children and their families for investing their time in the Danish High Risk and Resilience study. We thank Simon Yamazaki Jensen for assistance with statistical analyses, Line Carmichael for coordinating recruitment, and Jessica Ohland for handling the database. We also thank Agnete Albertsen, Anna Møller, Benthe Vink, Daban Sulaiman, Gøkze Akkas, Jonas Ingerslev, Malte Lundby, and Natascha Larsen at the DRCMR and Anette Bundgaard, Henriette Stadsgaard, Merete Birk, Nanna Steffensen, and Oskar Jefsen at CFIN for their effort in collecting the MRI data. The authors thank Daniel Gallichan for providing access to the FatNavs-MP2RAGE sequence. We used OpenAI’s ChatGPT (v4) for language editing, including refining sentence structure and shortening sections to improve clarity. This assistance was limited to text improvement and did not influence the scientific content or conclusions.

## Author Contribution

KSM had full access to all the data in the study and takes responsibility for the integrity of the data and the accuracy of the data analysis. MN, AAET, OM, LØ, and HRS designed the VIA study and applied for funding. KSM, MN, and HRS conceptualized the current study. JMB, MFK, MG, NH, AS, CBK, AKA, and LV recruited participants for the study and did the assessments of clinical symptoms. AAET supervised the clinical diagnosis. HS, TEL, and LØ were responsible for MR scanning. LKJ, KML, CBK, AKA, and LV scanned the participants. EH-T, WFCB, and KSM analyzed the MR data and created the figures. KSM, WFCB, and MN drafted the manuscript. All authors read and commented on the manuscript.

