## Supplementary material for "Sex-specific cortical brain differences in children at familial high risk for schizophrenia or bipolar disorder"

### Content

**Supplementary Results**

**Supplementary Discussion**

**Supplementary Figures**

Figure S1. Flowchart of the inclusion and exclusion procedures for the present structural MRI study.

Figure S2. Effect size map displaying the group difference in cortical thickness between children at familial high risk (FHR) for schizophrenia (FHR-SZ), bipolar disorder (FHR-BP), and population-based controls (PBC).

Figure S3. Effect size bar plots of the group difference in cortical volume between children at familial high risk (FHR) for schizophrenia (FHR-SZ) or bipolar disorder (FHR-BP) and population-based controls (PBC).

Figure S4. Effect size bar plots of the group difference in surface area between children at familial high risk (FHR) for schizophrenia (FHR-SZ) or bipolar disorder (FHR-BP) and population-based controls (PBC).

Figure S5. Effect size bar plots of the group difference in cortical thickness between children at familial high risk (FHR) for schizophrenia (FHR-SZ) or bipolar disorder (FHR-BP) and population-based controls (PBC).

**Supplementary Tables**

Table S1. Results from the drop-out analysis comparing VIA11 variables at age 11 years from participants included vs. not included in the current MRI study for each group.

Table S2. Results from the drop-out analysis comparing VIA7 variables at age 7 years from participants included vs. not included in the current MRI study for each group.

Table S3. Results from the primary analysis of group-by-sex effects on the global brain measures.

Table S4. Results from the analysis of group effects on the global brain measures.

Table S5. Results from the follow-up analysis of the global brain measures, controlling for height.

Table S6. Results from the follow-up analysis of the global brain measures, controlling for intracranial volume.

Table S7. Results from the pairwise group analysis of the global brain measures in males and females.

Table S8. Results from the pairwise group analysis of the global brain measures in males and females, controlling for axis-I diagnosis.

Table S9. Results from the pairwise analysis comparing participants *with* and *without* an axis-I diagnosis in each group.

### Supplementary Results

**Group-by-sex interaction and group effects on intracranial volume (ICV)**

Follow-up analysis revealed a significant group-by-sex interaction on ICV ($F = 5.293$, $p = 0.006$, $q = 0.015$, $BF_{10} = 7.908$), with the Bayes factor providing moderate evidence for a sex-specific group difference (Table S3). This group-by-sex interaction remained significant after adjusting for height as a proxy for general growth (Table S5). While height was significantly associated with ICV ($F = 18.034$, $p < 0.001$), it was not significantly associated with other global brain measures ($F < 3.760$, $p > 0.054$, Table S5). When ICV was included as a covariate in the models of brain volume, cortical volume, or surface area, none of the observed group-by-sex interactions remained significant ($p > 0.565$, Table S6). However, ICV was highly correlated with the global brain measures ($r$ range = 0.875 - 0.948, $p < 0.001$).

To facilitate comparison with existing literature, we conducted a post hoc analysis examining the main effect of group, excluding the group-by-sex interaction term. This analysis revealed a significant group effect ($F = 3.883$, $p = 0.022$, $BF_{10} = 1.405$), though the Bayes factor provided only anecdotal evidence (Table S4). Notably, the group effect was not significant when the group-by-sex interaction was included in the model ($F = 1.895$, $p = 0.152$, Table S3), indicating that sex-specific group differences were driving the overall effect.

**Group differences in ICV in males and females**

To disentangle the observed group-by-sex effects on ICV, we conducted pairwise group analyses for females and males separately (Table S7). Males at FHR-SZ displayed significantly smaller ICV than both FHR-BP and PBC males, with strong evidence ($BF_{10} > 10$) for the FHR-SZ vs. PBC comparison and very strong evidence ($BF_{10} > 30$) for the FHR-SZ vs. FHR-BP comparison. We did not observe a significant difference between FHR-BP and PBC males, with no evidence for a group difference ($BF_{10} = 0.249$). In females, we did not observe a significant group difference for ICV, and the Bayes factor indicated no evidence for group differences ($BF_{10} < 1.038$).

**Group differences in ICV in males and females: role of lifetime axis-I diagnosis**

We examined whether the group differences in ICV were related to an axis-I diagnosis. When additionally controlling for axis-I disorder, FHR-SZ males continued to exhibit significantly smaller ICV than FHR-BP and PBC males ($p < 0.009$, Table S8), suggesting the smaller ICV in FHR-SZ males was not mediated by those with an axis-I disorder. In females, no significant group differences in ICV emerged after controlling for axis-I diagnosis ($p > 0.116$, Table S8). Next, we stratified the groups by lifetime axis-I diagnosis status to explore whether children *with* (+) an axis-I diagnosis differed from those *without* (-). In males, we did not observe significant differences between individuals with or without an axis-I diagnosis within any groups ($p > 0.339$, Table S9), suggesting that axis-I diagnosis was not associated with ICV variation. In females, FHR-SZ+ individuals displayed significantly larger ICV than FHR-SZ- females ($t = 2.399$, $p = 0.021$), while no significant differences were observed in the FHR-BP or PBC groups ($p > 0.279$, Table S9).

**Group differences in cortical thickness across the cortex in males and females**

Parcel-based analyses were conducted to explore potential group differences in cortical thickness across the cortex in males and females. The effect sizes obtained from these analyses are visualized on brain maps in Figure S2 and bar plots in Figure S5. The effect sizes were generally modest, and only a few regions displayed at least medium effect size (Cohen's D above 0.5, ranging between 0.50-0.72). FHR-BP females displayed thicker cortex in the right supramarginal, inferior parietal, and precentral cortices than PBC females, while FHR-BP males had thinner cortex in the right isthmus and the cingulate cortex than the PBC males. Moreover, FHR-SZ females had thinner right paracentral and rostral middle frontal cortices than FHR-BP females, and FHR-SZ males displayed thinner cortex in the right middle and superior temporal as well as the left pars orbitalis and middle temporal cortices. None of the regions survived FDR correction.

### Supplementary Discussion

We observed significant group-by-sex differences in ICV. Males at FHR-SZ displayed smaller ICV than FHR-BP and PBC males, with the Bayes factor providing strong evidence for these associations. In females, we did not observe significant effects or evidence for group differences in ICV.

The group-by-sex effects in brain volume, cortical volume, and surface area did not remain significant after adjusting for ICV. ICV demonstrated very strong positive correlations with total brain volume, cortical volume, and surface area, which is characteristic in pediatric cohorts due to the close relationship between brain growth and skull growth during development (1–3). Consequently, it is challenging to separate ICV from global brain measures in these populations. Importantly, the group-by-sex effects persisted when controlling for height, suggesting that the observed associations were not driven by individual differences in overall growth.

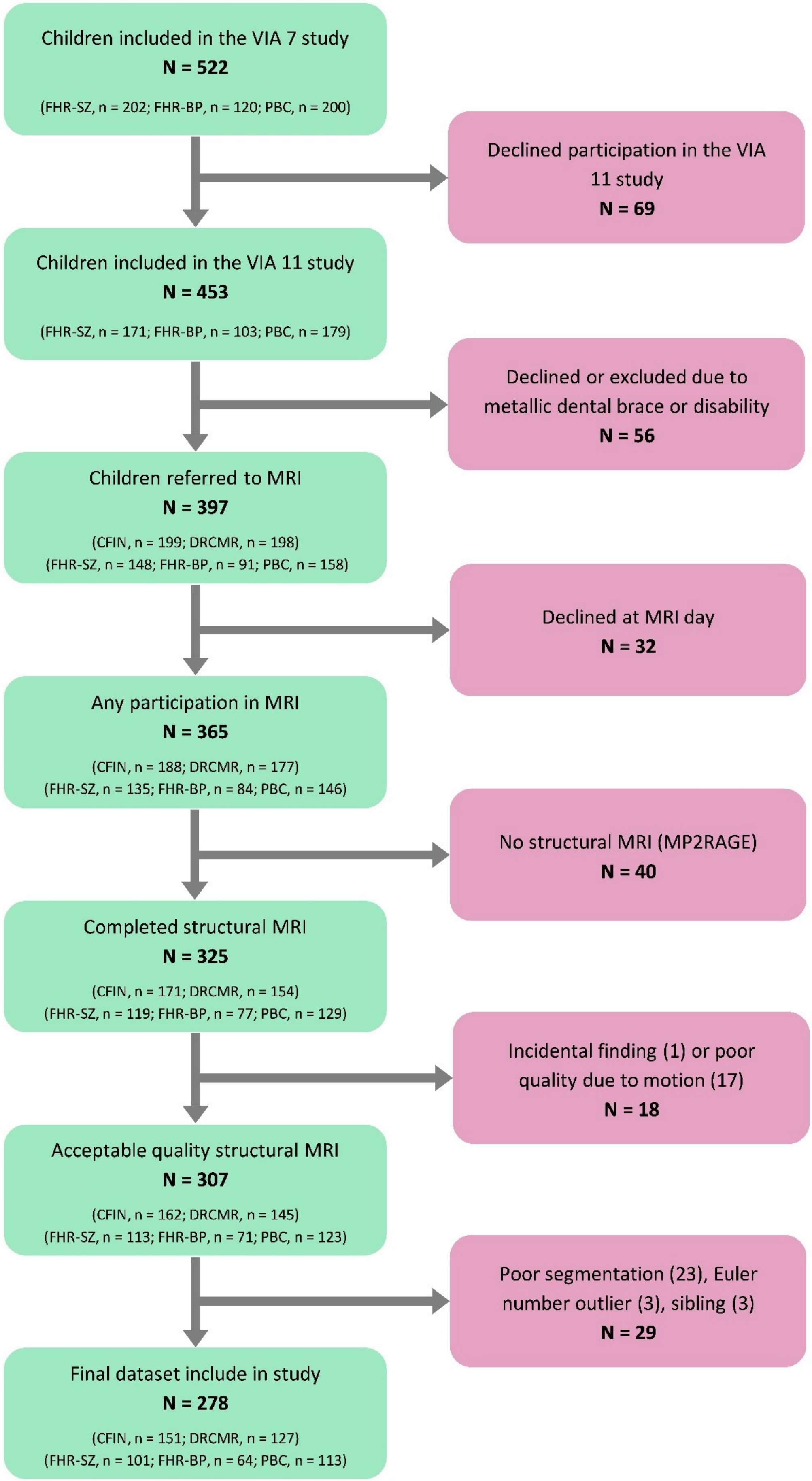

Figure S1. Flowchart of the inclusion and exclusion procedures for the present structural MRI study. Abbreviations: CFIN: Center of Functionally Integrative Neuroscience; DRCMR: Danish Research Center for Magnetic Resonance; FHR-SZ: familial high risk of schizophrenia; FHR-BP: familial high risk of bipolar disorder; PBC: population-based controls.

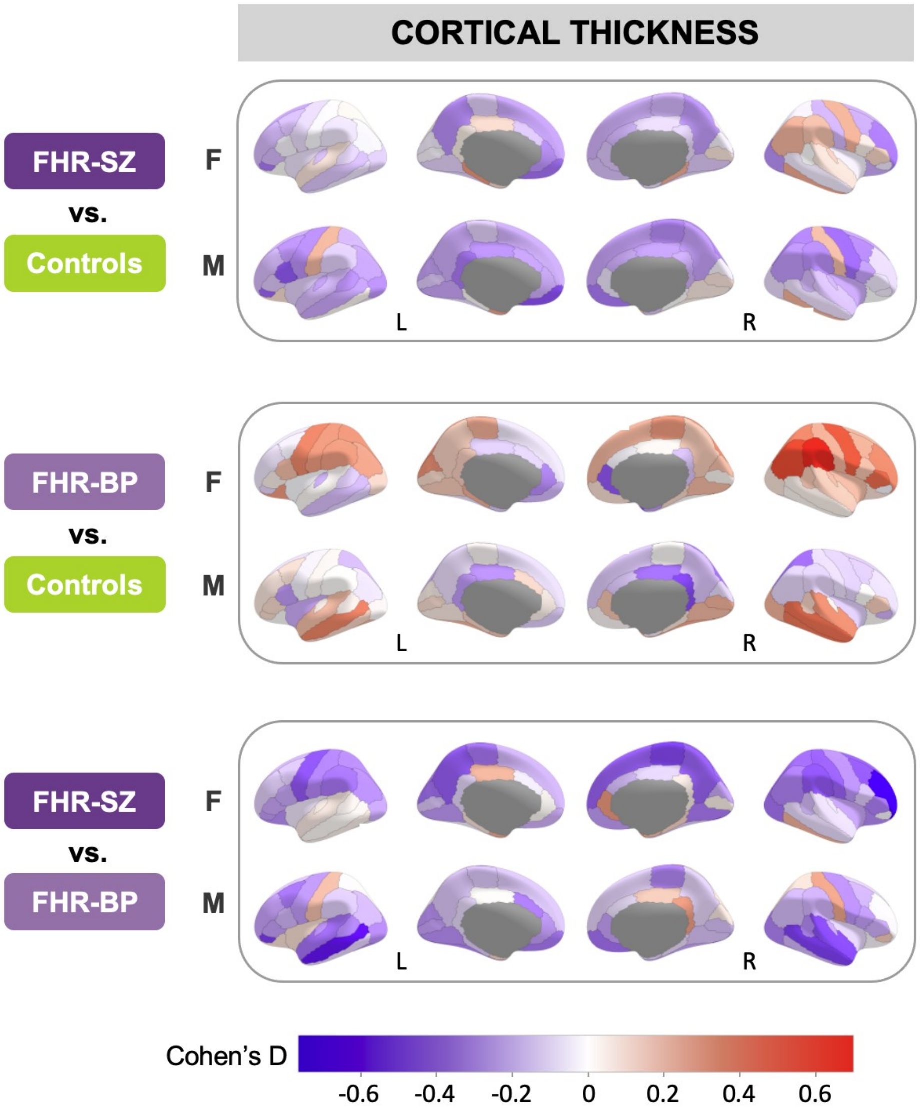

Figure S2. Effect size map displaying the group difference in cortical thickness between children at familial high risk for schizophrenia (FHR-SZ, top) or bipolar disorder (FHR-BP, middle) vs. population-based controls and between FHR-SZ vs. FHR-BP (bottom) for females (F) and males (M). Red shades indicate regions where FHR groups or FHR-SZ have larger cortical volume or surface area than, respectively, controls or FHR-BP. Blue shades indicate regions where FHR groups or FHR-SZ have smaller cortical volume or surface area than controls or FHR-BP. None of the parcels were significant after FDR correction.

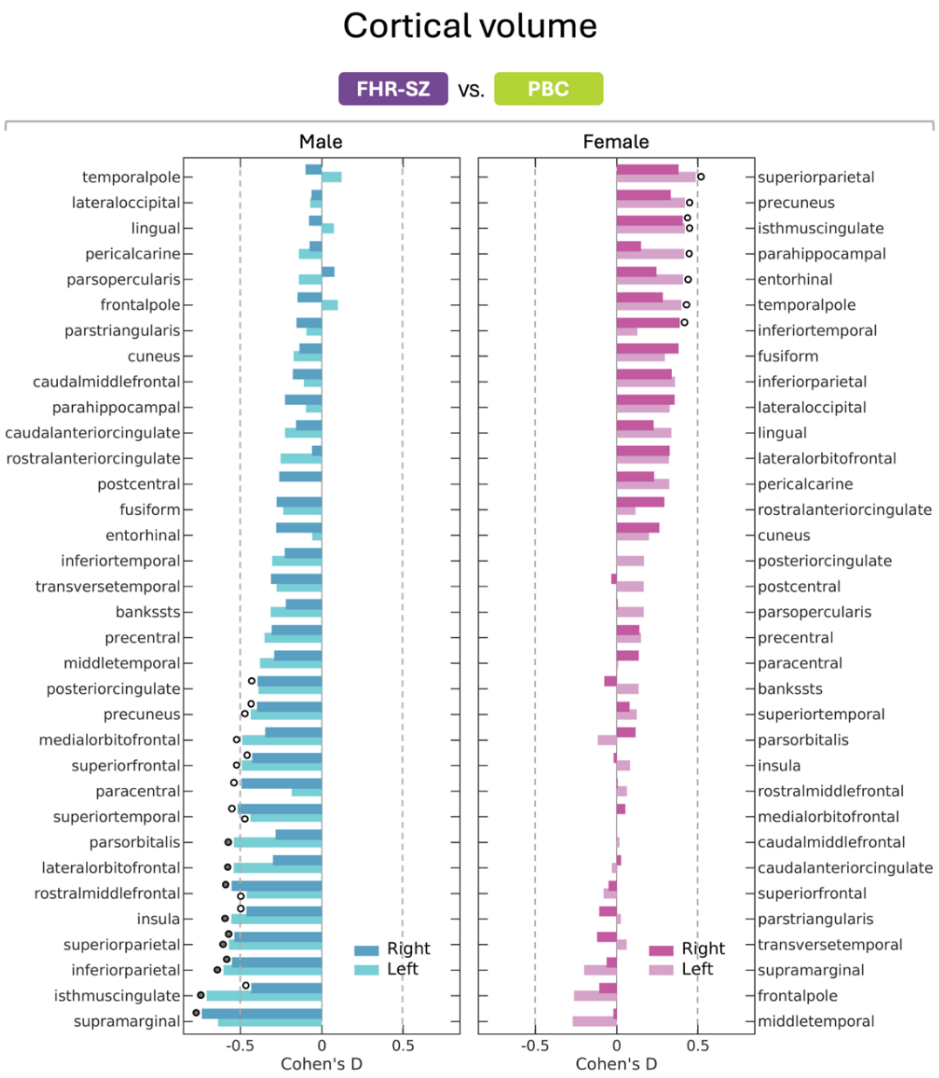
Cortical volume
FHR-SZ vs. PBC
Male
Female
temporalpole
lateraloccipital
lingual
pericalcarine
parsopercularis
frontalpole
parstriangularis
cuneus
caudalmiddlefrontal
parahippocampal
caudalanteriorcingulate
rostralanteriorcingulate
postcentral
fusiform
entorhinal
inferiortemporal
transversetemporal
bankssts
precentral
middletemporal
posteriorcingulate
precuneus
medialorbitofrontal
superiorfrontal
paracentral
superiortemporal
parsorbitalis
lateralorbitofrontal
rostralmiddlefrontal
insula
superiorparietal
inferiorparietal
isthmuscingulate
supramarginal
superiorparietal
precuneus
isthmuscingulate
parahippocampal
entorhinal
temporalpole
inferiortemporal
fusiform
inferiorparietal
lateraloccipital
lingual
lateralorbitofrontal
pericalcarine
rostralanteriorcingulate
cuneus
posteriorcingulate
postcentral
parsopercularis
precentral
paracentral
bankssts
superiortemporal
parsorbitalis
insula
rostralmiddlefrontal
medialorbitofrontal
caudalmiddlefrontal
caudalanteriorcingulate
superiorfrontal
parstriangularis
transversetemporal
supramarginal
frontalpole
middletemporal
Right
Left
Right
Left
-0.5
0
0.5
Cohen's D
-0.5
0
0.5
Cohen's D

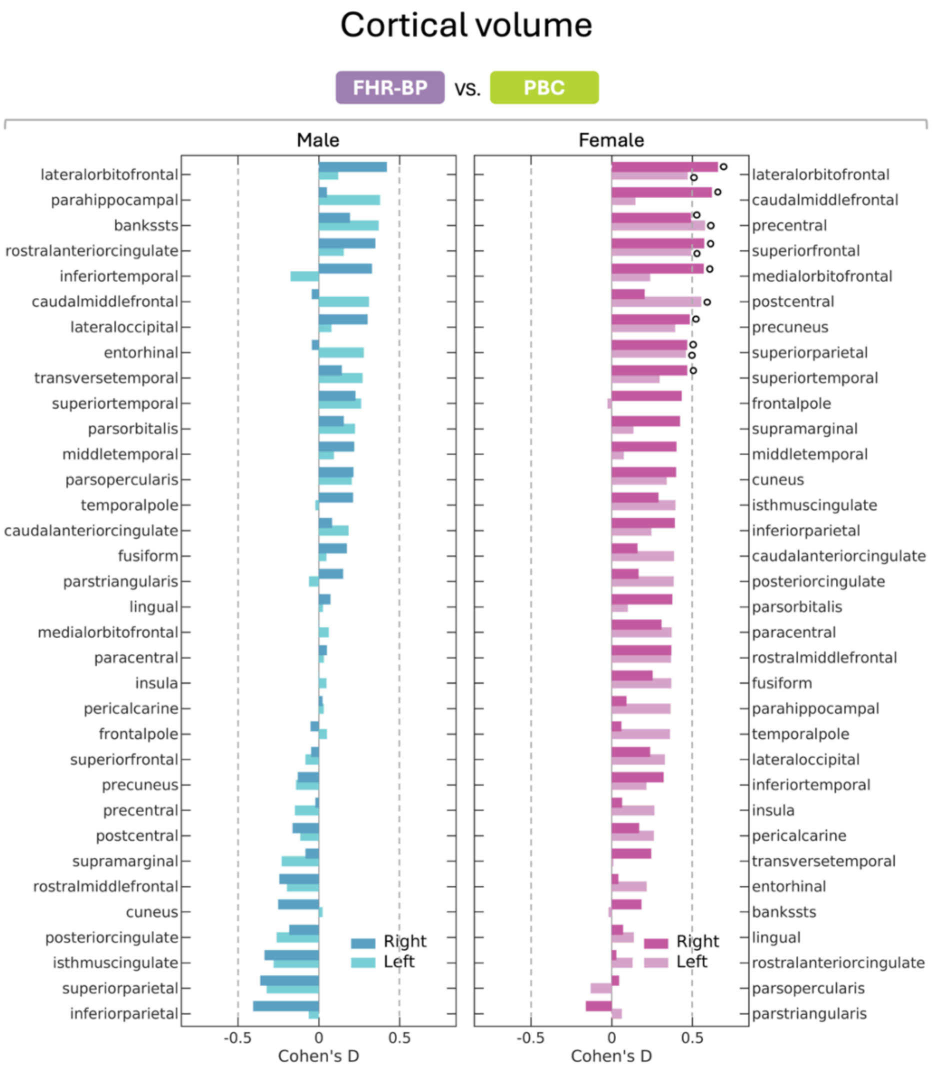
Cortical volume
FHR-BP vs. PBC
Male
Female
lateralorbitofrontal
parahippocampal
bankssts
rostralanteriorcingulate
inferiortemporal
caudalmiddlefrontal
lateraloccipital
entorhinal
transversetemporal
superiortemporal
parsorbitalis
middletemporal
parsopercularis
temporalpole
caudalanteriorcingulate
fusiform
parstriangularis
lingual
medialorbitofrontal
paracentral
insula
pericalcarine
frontalpole
superiorfrontal
precuneus
precentral
postcentral
supramarginal
rostralmiddlefrontal
cuneus
posteriorcingulate
isthmuscingulate
superiorparietal
inferiorparietal
lateralorbitofrontal
caudalmiddlefrontal
precentral
superiorfrontal
medialorbitofrontal
postcentral
precuneus
superiorparietal
superiortemporal
frontalpole
supramarginal
middletemporal
cuneus
isthmuscingulate
inferiorparietal
caudalanteriorcingulate
posteriorcingulate
parsorbitalis
paracentral
rostralmiddlefrontal
fusiform
parahippocampal
temporalpole
lateraloccipital
inferiortemporal
insula
pericalcarine
transversetemporal
entorhinal
bankssts
lingual
rostralanteriorcingulate
parsopercularis
parstriangularis
Right
Left
Right
Left
-0.5
0
0.5
Cohen's D
-0.5
0
0.5
Cohen's D

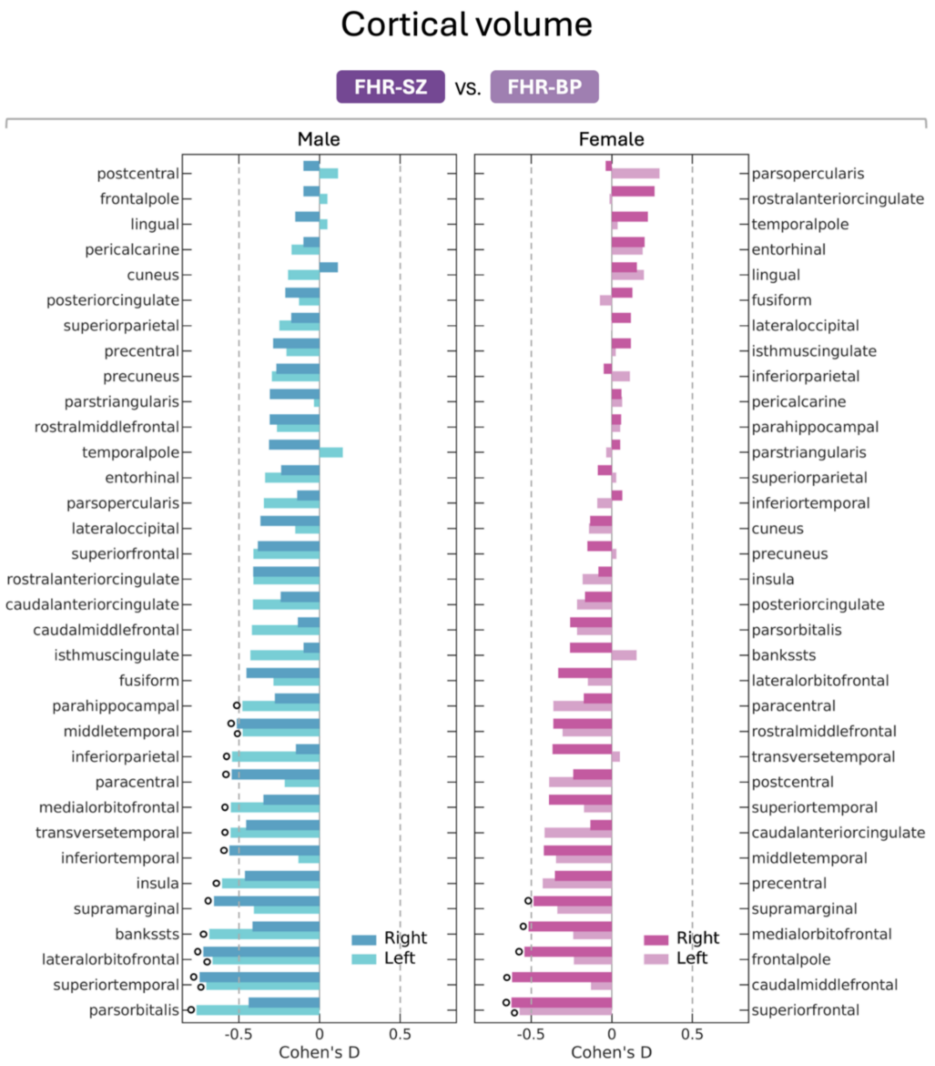

Figure S3. Effect size bar plots of the group difference in cortical volume between children at familial high risk (FHR) for schizophrenia (FHR-SZ, top left) or bipolar disorder (FHR-BP, top right), and population-based controls (PBC, bottom). Regions are ranked from the highest to the lowest Cohen's D value in either hemisphere. Regions significant at an uncorrected p-value of 0.05 are indicated by an open circle (○) next to the bar. Regions that survive FDR correction (q ≤ 0.05) are indicated with a closed circle (●).

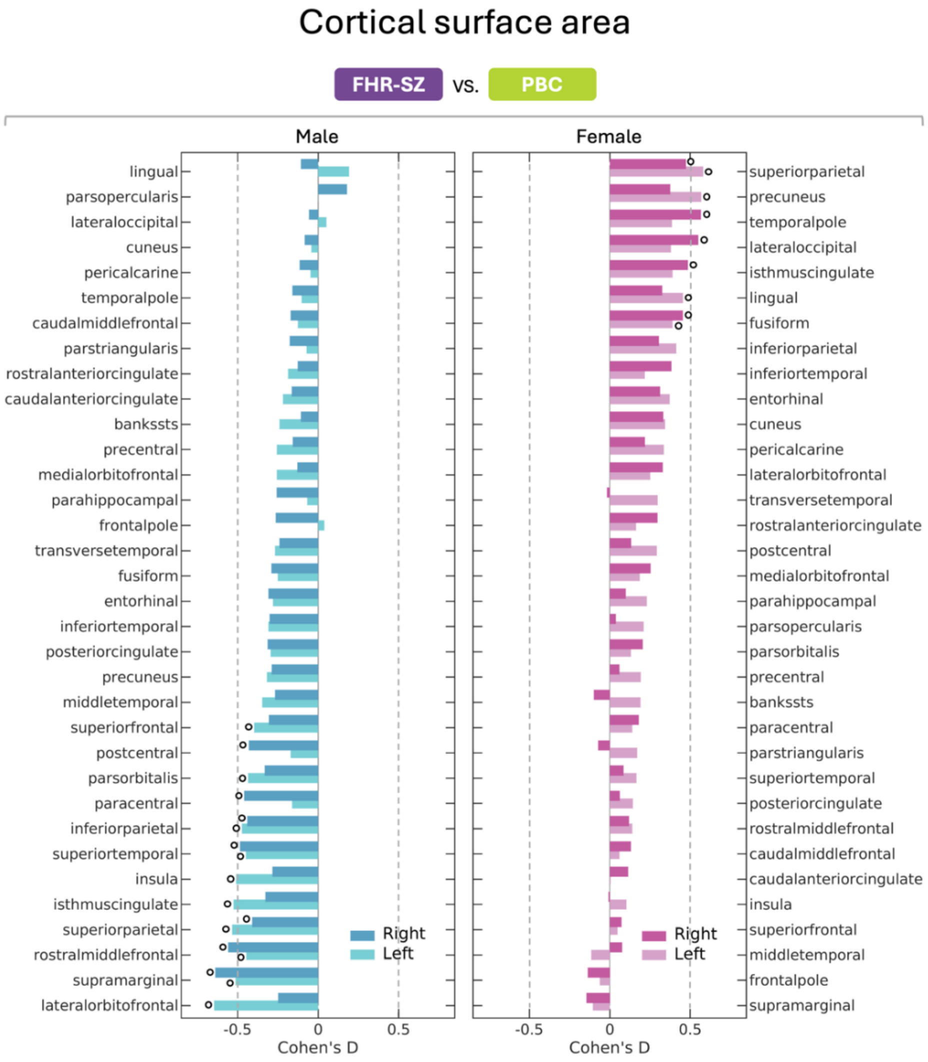
Cortical surface area
FHR-SZ vs. PBC
Male
Female
lingual
parsopercularis
lateraloccipital
cuneus
pericalcarine
temporalpole
caudalmiddlefrontal
parstriangularis
rostralanteriorcingulate
caudalanteriorcingulate
bankssts
precentral
medialorbitofrontal
parahippocampal
frontalpole
transversetemporal
fusiform
entorhinal
inferiortemporal
posteriorcingulate
precuneus
middletemporal
superiorfrontal
postcentral
parsorbitalis
paracentral
inferiorparietal
superiortemporal
insula
isthmuscingulate
superiorparietal
rostralmiddlefrontal
supramarginal
lateralorbitofrontal
superiorparietal
precuneus
temporalpole
lateraloccipital
isthmuscingulate
lingual
fusiform
inferiorparietal
inferiortemporal
entorhinal
cuneus
pericalcarine
lateralorbitofrontal
transversetemporal
rostralanteriorcingulate
postcentral
medialorbitofrontal
parahippocampal
parsopercularis
parsorbitalis
precentral
bankssts
paracentral
parstriangularis
superiortemporal
posteriorcingulate
rostralmiddlefrontal
caudalmiddlefrontal
caudalanteriorcingulate
insula
superiorfrontal
middletemporal
frontalpole
supramarginal
Right
Left
-0.5
0
0.5
Cohen's D

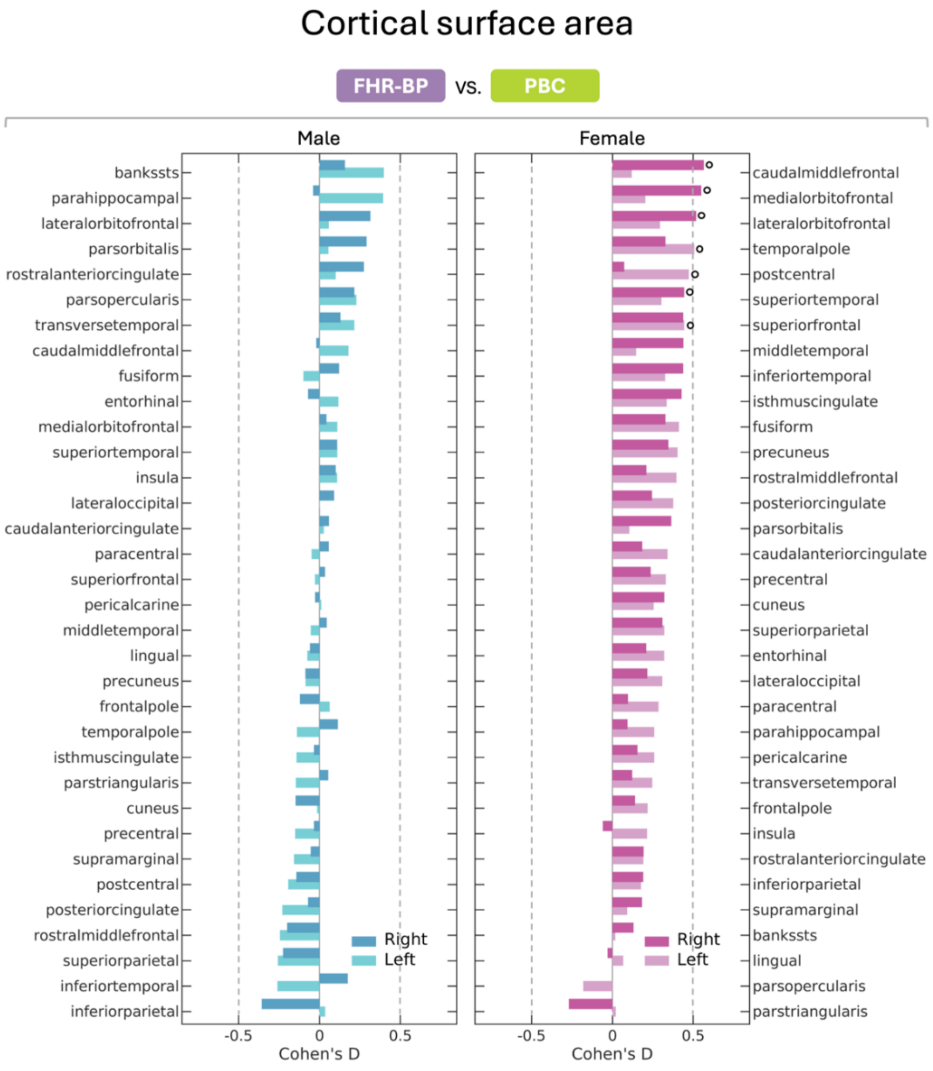
Cortical surface area
FHR-BP vs. PBC
Male
Female
bankssts
parahippocampal
lateralorbitofrontal
parsorbitalis
rostralanteriorcingulate
parsopercularis
transversetemporal
caudalmiddlefrontal
fusiform
entorhinal
medialorbitofrontal
superiortemporal
insula
lateraloccipital
caudalanteriorcingulate
paracentral
superiorfrontal
pericalcarine
middletemporal
lingual
precuneus
frontalpole
temporalpole
isthmuscingulate
parstriangularis
cuneus
precentral
supramarginal
postcentral
posteriorcingulate
rostralmiddlefrontal
superiorparietal
inferiortemporal
inferiorparietal
caudalmiddlefrontal
medialorbitofrontal
lateralorbitofrontal
temporalpole
postcentral
superiortemporal
superiorfrontal
middletemporal
inferiortemporal
isthmuscingulate
fusiform
precuneus
rostralmiddlefrontal
posteriorcingulate
parsorbitalis
caudalanteriorcingulate
precentral
cuneus
superiorparietal
entorhinal
lateraloccipital
paracentral
parahippocampal
pericalcarine
transversetemporal
frontalpole
insula
rostralanteriorcingulate
inferiorparietal
supramarginal
bankssts
lingual
parsopercularis
parstriangularis
Right
Left
-0.5
0
0.5
Cohen's D

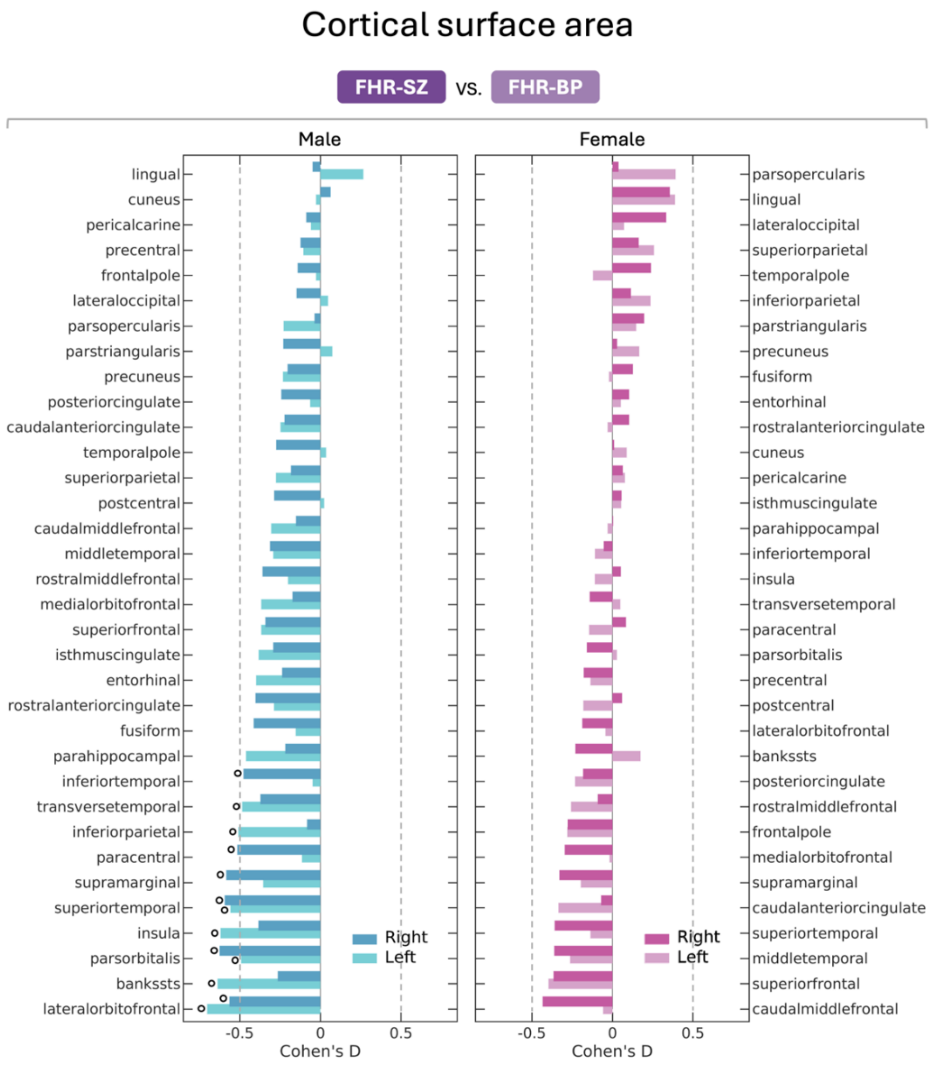

Figure S4. Effect size bar plots of the group difference in surface area between children at familial high risk (FHR) for schizophrenia (FHR-SZ, top left) or bipolar disorder (FHR-BP, top right), and population-based controls (PBC, bottom). Regions are ranked from the highest to the lowest Cohen's D value in either hemisphere. Regions significant at an uncorrected p-value of 0.05 are indicated by an open circle (○) next to the bar. Regions that survive FDR correction (q ≤ 0.05) are indicated with a closed circle (●).

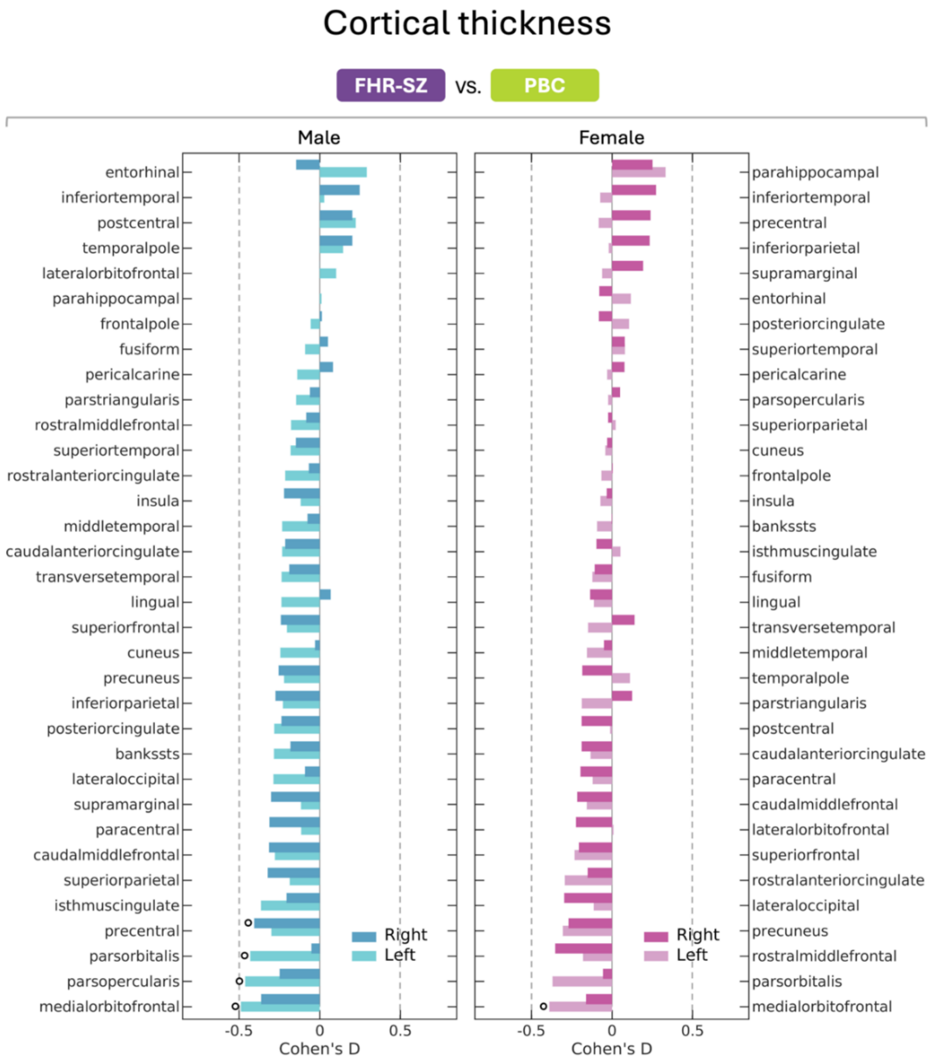

Cortical thickness
FHR-SZ vs. PBC
Male
Female
entorhinal
inferiortemporal
postcentral
temporalpole
lateralorbitofrontal
parahippocampal
frontalpole
fusiform
pericalcarine
parstriangularis
rostralmiddlefrontal
superiortemporal
rostralanteriorcingulate
insula
middletemporal
caudalanteriorcingulate
transversetemporal
lingual
superiorfrontal
cuneus
precuneus
inferiorparietal
posteriorcingulate
bankssts
lateraloccipital
supramarginal
paracentral
caudalmiddlefrontal
superiorparietal
isthmuscingulate
precentral
parsorbitalis
parsopercularis
medialorbitofrontal
parahippocampal
inferiortemporal
precentral
inferiorparietal
supramarginal
entorhinal
posteriorcingulate
superiortemporal
pericalcarine
parsopercularis
superiorparietal
cuneus
frontalpole
insula
bankssts
isthmuscingulate
fusiform
lingual
transversetemporal
middletemporal
temporalpole
parstriangularis
postcentral
caudalanteriorcingulate
paracentral
caudalmiddlefrontal
lateralorbitofrontal
superiorfrontal
rostralanteriorcingulate
lateraloccipital
precuneus
rostralmiddlefrontal
parsorbitalis
medialorbitofrontal
Right
Left
-0.5
0
0.5
Cohen's D

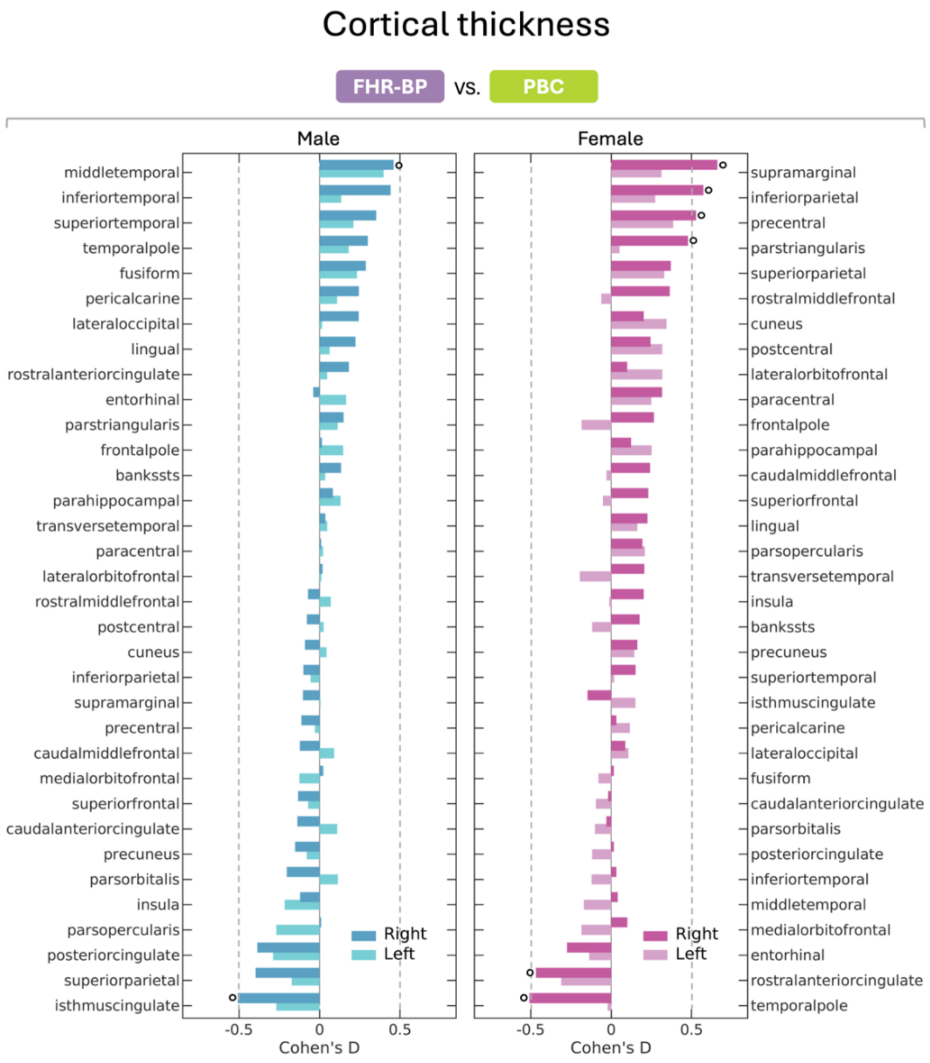

Cortical thickness
FHR-BP vs. PBC
Male
Female
middletemporal
inferiortemporal
superiortemporal
temporalpole
fusiform
pericalcarine
lateraloccipital
lingual
rostralanteriorcingulate
entorhinal
parstriangularis
frontalpole
bankssts
parahippocampal
transversetemporal
paracentral
lateralorbitofrontal
rostralmiddlefrontal
postcentral
cuneus
inferiorparietal
supramarginal
precentral
caudalmiddlefrontal
medialorbitofrontal
superiorfrontal
caudalanteriorcingulate
precuneus
parsorbitalis
insula
parsopercularis
posteriorcingulate
superiorparietal
isthmuscingulate
supramarginal
inferiorparietal
precentral
parstriangularis
superiorparietal
rostralmiddlefrontal
cuneus
postcentral
lateralorbitofrontal
paracentral
frontalpole
parahippocampal
caudalmiddlefrontal
superiorfrontal
lingual
parsopercularis
transversetemporal
insula
bankssts
precuneus
superiortemporal
isthmuscingulate
pericalcarine
lateraloccipital
fusiform
caudalanteriorcingulate
parsorbitalis
posteriorcingulate
inferiortemporal
middletemporal
medialorbitofrontal
entorhinal
rostralanteriorcingulate
temporalpole
Right
Left
-0.5
0
0.5
Cohen's D

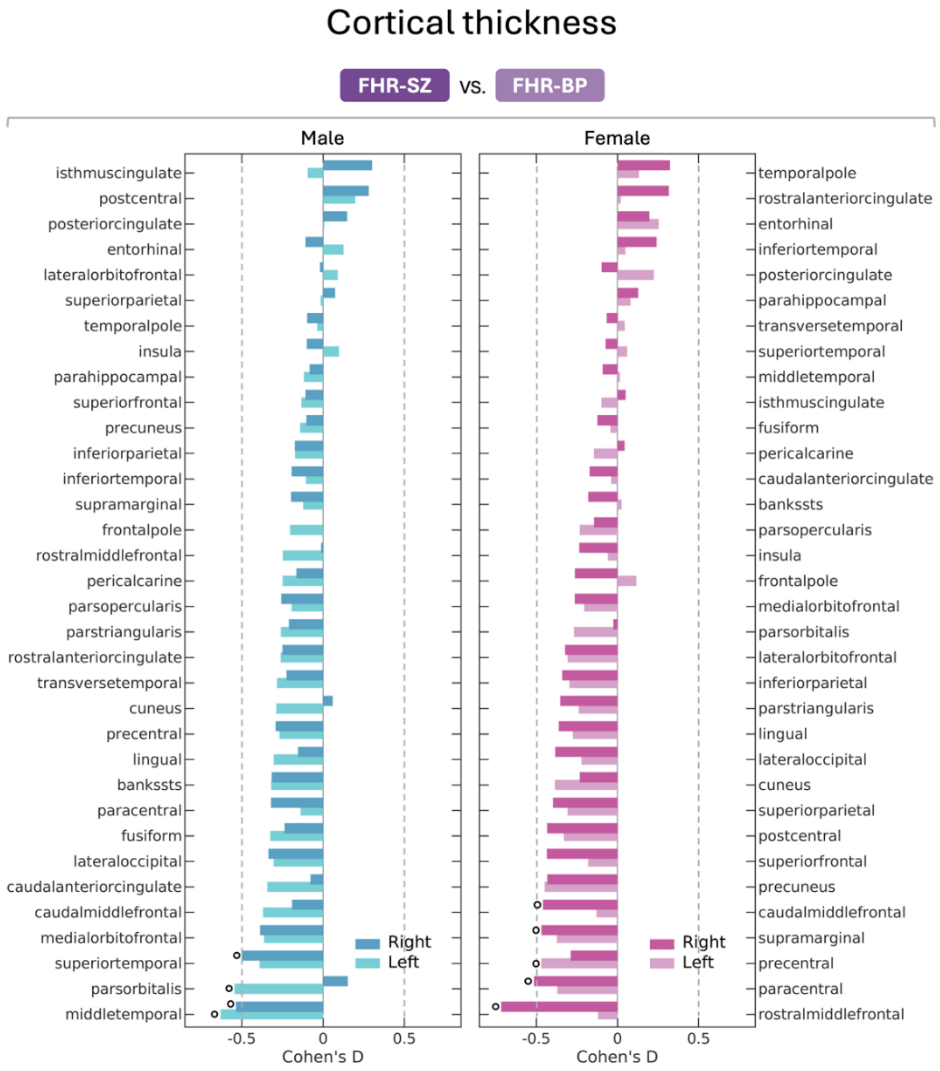

Figure S5. Effect size bar plots of the group difference in cortical thickness between children at familial high risk (FHR) for schizophrenia (FHR-SZ, top left) or bipolar disorder (FHR-BP, top right), and population-based controls (PBC, bottom). Regions are ranked from the highest to the lowest Cohen's D value in either hemisphere. Regions significant at an uncorrected p-value of 0.05 are indicated by an open circle (○) next to the bar. Regions that survive FDR correction (q ≤ 0.05) are indicated with a closed circle (●).

**Table S1. Results from the drop-out analysis comparing VIA11 variables at age 11 years from participants included vs. not included in the current MRI study for each group.**

| | FHR-SZ | | | FHR-BP | | | PBC | | |
|---|---|---|---|---|---|---|---|---|---|
| **VIA11** | Not included | Included | p | Not included | Included | p | Not included | Included | p |
| Children, N | 70 | 101 | | 39 | 64 | | 66 | 113 | |
| Females, N (%) | 32 (45.7%) | 50 (49.5%) | 0.538 | 13 (33.3%) | 32 (50.0%) | 0.110 | 27 (40.9%) | 56 (49.6%) | 0.198 |
| CBCL Total, mean (SD)* | 27.2 (23.6) | 20.9 (17.9) | **0.028** | 22.1 (21.3) | 20.9 (21.2) | 0.733 | 14.2 (15.1) | 11.6 (10.7) | 0.372 |
| CGAS, mean (SD)* | 59.9 (15.8) | 67.8 (14.7) | **<0.001** | 66.0 (14.2) | 69.5 (15.4) | 0.244 | 74.0 (13.5) | 75.9 (14.2) | 0.407 |
| Any axis-I diagnosis, N (%)* | 41 (58.6%) | 51 (51.5%) | 0.377 | 23 (59.0%) | 31 (48.4%) | 0.250 | 19 (29.7%) | 31 (27.9%) | 0.804 |
| Primary caregiver's PSP, mean (SD)* | 66.6 (17.1) | 72.5 (16.5) | **0.010** | 72.2 (15.6) | 71.5 (15.5) | 0.824 | 84.6 (7.98) | 82.6 (11.2) | 0.368 |

Drop-out analysis tested for differences in the VIA11 data between participants included vs. non-included in the present study for each group (FHR-SZ, FHR-BP, PBC). Two-tailed t-tests were used to assess CBCL, CGAS, and PSP. A chi-square test assessed sex and prevalence of lifetime axis-I disorders. Significant p-values are shown in bold. Abbreviations: FHR-SZ: Familial high risk for schizophrenia, FHR-BP: Familial high risk for bipolar disorder, PBC: Population-based controls, CBCL: Child Behavior Check List, CGAS: Children's Global Assessment Scale, PSP: Personal and Social Performance. *Missing data in VIA11: CBCL (n=18), CGAS (n=4), axis-I disorders (n=6), PSP (n=7).

**Table S2. Results from the drop-out analysis comparing VIA7 variables at age 7 years from participants included vs. not included in the current MRI study for each group.**

| | FHR-SZ | | | FHR-BP | | | PBC | | |
|---|---|---|---|---|---|---|---|---|---|
| **VIA7** | Not included | Included | p | Not included | Included | p | Not included | Included | p |
| Children, N | 101 | 101 | | 56 | 64 | | 87 | 113 | |
| Females, N (%) | 43 (42.6%) | 50 (49.5%) | 0.323 | 24 (42.9%) | 32 (50.0%) | 0.434 | 37 (42.5%) | 56 (49.6%) | 0.323 |
| CBCL Total, mean (SD)* | 28.5 (23.1) | 26.0 (18.9) | 0.351 | 23.7 (19.7) | 23.2 (19.8) | 0.883 | 18.5 (15.5) | 15.8 (14.0) | 0.318 |
| CGAS, mean (SD)* | 64.8 (15.2) | 71.4 (15.0) | **0.001** | 72.6 (15.6) | 74.4 (14.4) | 0.492 | 76.5 (13.9) | 78.6 (13.1) | 0.328 |
| Any axis-I diagnosis, N (%)* | 45 (44.6%) | 32 (31.7%) | 0.066 | 21 (37.5%) | 21 (32.8%) | 0.492 | 13 (14.9%) | 17 (15.0%) | 0.933 |
| Primary caregiver's PSP, mean (SD)* | 70.5 (12.6) | 75.8 (15.1) | **0.003** | 73.4 (14.4) | 75.3 (14.1) | 0.422 | 84.3 (9.36) | 84.5 (9.03) | 0.884 |

Drop-out analysis tested for differences in the VIA7 data between participants included vs. non-included in the present study for each group (FHR-SZ, FHR-BP, PBC). Two-tailed t-tests were used to assess CBCL, CGAS, and PSP. Chi-square tests assessed sex and prevalence of lifetime axis-I disorders. Significant p-values are shown in bold. Abbreviations: FHR-SZ: Familial high risk for schizophrenia, FHR-BP: Familial high risk for bipolar disorder, PBC: Population-based controls, CBCL: Child Behavior Check List, CGAS: Children's Global Assessment Scale, PSP: Personal and Social Performance. *Missing data: CBCL (n=28), CGAS (n=8), axis-I disorders (n=8), PSP (n=10).

**Table S3. Results from the primary analysis of group-by-sex effects on the global brain measures.**

| | Group | | | | Group-by-sex | | | | Age | | Sex | | MR-site | | Euler number | |
|---|---|---|---|---|---|---|---|---|---|---|---|---|---|---|---|---|
| | $Eta^2$ | $BF_{10}$ | F | p | $Eta^2$ | $BF_{10}$ | F | p | F | p | F | p | F | p | F | p |
| Brain volume | 0.018 | 16.066 | 2.525 | 0.082 | 0.038 | 8.362 | 5.270 | **0.006** | 0.534 | 0.466 | 18.691 | <0.001 | 7.348 | 0.007 | 9.002 | 0.003 |
| Cortical volume | 0.019 | 4.731 | 2.596 | 0.076 | 0.031 | 3.523 | 4.364 | **0.014** | 1.741 | 0.188 | 12.068 | <0.001 | 3.144 | 0.077 | 0.912 | 0.340 |
| Surface area | 0.015 | 0.457 | 2.063 | 0.129 | 0.030 | 2.982 | 4.220 | **0.016** | 1.306 | 0.254 | 17.220 | <0.001 | 0.467 | 0.495 | 0.143 | 0.706 |
| Cortical thickness | 0.020 | 0.044 | 1.571 | 0.210 | 0.002 | 0.088 | 0.205 | 0.815 | 0.497 | 0.481 | 0.522 | 0.471 | 3.653 | 0.057 | 12.259 | <0.001 |
| Intracranial volume | 0.014 | 11.368 | 1.895 | 0.152 | 0.038 | 7.908 | 5.293 | **0.006** | 0.013 | 0.910 | 16.827 | <0.001 | 1.273 | 0.260 | 7.180 | 0.008 |

Each row represents a separate ANCOVA model, examining group-by-sex differences. All models included the covariates: age, sex, MR-site, and Euler number. Significant group-by-sex effects are shown in bold. All significant group-by-sex effects survived FDR correction. $Eta^2$ and Bayes factor ($BF_{10}$) are given for the group and group-by-sex effects. Number of male/female children in each group: FHR-SZ: n=51/50, FHR-BP: n=32/32, PBC: n=57/56.

**Table S4. Results from the analysis of group effects on the global brain measures.**

| | Group | | | | Age | | Sex | | MR-site | | Euler number | |
|---|---|---|---|---|---|---|---|---|---|---|---|---|
| | $Eta^2$ | $BF_{10}$ | F | p | F | p | F | p | F | p | F | p |
| Brain volume | 0.031 | 1.885 | 4.288 | **0.015** | 0.423 | 0.516 | 76.936 | <0.001 | 8.481 | 0.004 | 11.788 | <0.001 |
| Cortical volume | 0.028 | 1.498 | 3.937 | **0.021** | 1.364 | 0.244 | 63.643 | <0.001 | 3.774 | 0.053 | 1.676 | 0.197 |
| Surface area | 0.010 | 0.141 | 1.380 | 0.253 | 1.068 | 0.302 | 76.269 | <0.001 | 0.765 | 0.383 | 0.550 | 0.459 |
| Cortical thickness | 0.019 | 0.507 | 2.775 | 0.064 | 0.387 | 0.535 | 0.311 | 0.578 | 3.706 | 0.055 | 12.659 | <0.001 |
| Intracranial volume | 0.022 | 1.405 | 3.883 | **0.022** | 0.034 | 0.854 | 67.766 | <0.001 | 1.825 | 0.178 | 9.725 | 0.002 |

Each row represents a separate ANCOVA model, examining group differences. All models included the covariates: age, sex, MR-site, and Euler number. Significant group effects are shown in bold. All significant group effects survived FDR correction. $Eta^2$ and Bayes factor ($BF_{10}$) are given for the group effects. Number of male/female children in each group: FHR-SZ: n=51/50, FHR-BP: n=32/32, PBC: n=57/56.

**Table S5. Results from the follow-up analysis of the global brain measures, controlling for height.**

| | Group | | Group-by-sex | | Age | | Sex | | MR-site | | Euler number | | Height | |
|---|---|---|---|---|---|---|---|---|---|---|---|---|---|---|
| | F | p | F | p | F | p | F | p | F | p | F | p | F | p |
| Brain volume | 2.365 | 0.096 | 4.677 | **0.010** | 1.071 | 0.302 | 19.714 | <0.001 | 7.522 | 0.007 | 8.355 | 0.004 | 3.760 | 0.054 |
| Cortical volume | 2.487 | 0.085 | 4.019 | **0.019** | 2.190 | 0.140 | 12.447 | <0.001 | 3.183 | 0.076 | 0.784 | 0.377 | 1.224 | 0.270 |
| Surface area | 1.980 | 0.140 | 3.898 | **0.021** | 1.669 | 0.197 | 17.620 | <0.001 | 0.481 | 0.489 | 0.098 | 0.754 | 1.052 | 0.306 |
| Intracranial volume | 1.668 | 0.191 | 4.278 | **0.015** | 0.318 | 0.573 | 19.751 | <0.001 | 1.448 | 0.230 | 6.230 | 0.013 | 18.034 | <0.001 |

Each row represents a separate ANCOVA model, examining group-by-sex differences when controlling for height. All models included the covariates: age, sex, MR-site, Euler number, and height. Significant group-by-sex effects at the uncorrected 0.05 level are shown in bold. Number of male/female children in each group: FHR-SZ: n=51/50, FHR-BP: n=32/32, PBC: n=57/56.

**Table S6. Results from the follow-up analysis of the global brain measures, controlling for intracranial volume.**

| | Group | | Group-by-sex | | Age | | Sex | | MR-site | | Euler number | | ICV | |
|---|---|---|---|---|---|---|---|---|---|---|---|---|---|---|
| | F | p | F | p | F | p | F | p | F | p | F | p | F | p |
| Brain volume | 0.694 | 0.500 | 0.181 | 0.834 | 5.131 | 0.024 | 1.795 | 0.181 | 20.189 | <0.001 | 1.855 | 0.174 | 1708.7 | <0.001 |
| Cortical volume | 0.886 | 0.414 | 0.573 | 0.565 | 7.091 | 0.008 | 0.000 | 0.995 | 2.350 | 0.126 | 5.979 | 0.015 | 684.7 | <0.001 |
| Surface area | 5.878 | **0.016** | 0.190 | 0.827 | 6.837 | 0.009 | 1.207 | 0.273 | 0.424 | 0.516 | 16.952 | <0.001 | 924.3 | <0.001 |

Each row represents a separate ANCOVA model, examining group-by-sex differences when controlling for intracranial volume (ICV). All models included the covariates: age, sex, MR-site, and Euler number. Significant group effects at the uncorrected 0.05 level are shown in bold. Number of male/female children in each group: FHR-SZ: n=51/50, FHR-BP: n=32/32, PBC: n=57/56.

### Table S7. Results from the pairwise group analysis of the global brain measures in males and females.

| | LS means | | | FHR-SZ vs. PBC | | | | FHR-BP vs. PBC | | | | FHR-SZ vs. FHR-BP | | | |
|---|---|---|---|---|---|---|---|---|---|---|---|---|---|---|---|
| | FHR-SZ | FHR-BP | PBC | *d* | $BF_{10}$ | t | p | *d* | $BF_{10}$ | t | p | *d* | $BF_{10}$ | t | p |
| **Females** | | | | | | | | | | | | | | | |
| Brain volume | 1207948 | 1228397 | 1181961 | 0.294 | 0.535 | 1.500 | 0.136 | 0.525 | 1.858 | 2.353 | **0.020** | 0.231 | 0.401 | -1.014 | 0.312 |
| Cortical volume | 579347 | 592192 | 568917 | 0.241 | 0.387 | 1.230 | 0.221 | 0.537 | 2.062 | 2.408 | **0.017** | 0.297 | 0.546 | -1.301 | 0.196 |
| Surface area | 183275 | 184232 | 178492 | 0.336 | 0.701 | 1.715 | 0.089 | 0.403 | 0.760 | 1.807 | 0.073 | 0.067 | 0.263 | -0.295 | 0.769 |
| Intracranial volume | 1460212 | 1480260 | 1435861 | 0.236 | 0.371 | 1.207 | 0.229 | 0.431 | 1.038 | 1.932 | 0.056 | 0.195 | 0.340 | -0.854 | 0.395 |
| **Males** | | | | | | | | | | | | | | | |
| Brain volume | 1255717 | 1320529 | 1312553 | -0.624 | 15.368 | -3.157 | **0.002** | 0.087 | 0.279 | 0.385 | 0.701 | 0.711 | 20.623 | -3.026 | **0.003** |
| Cortical volume | 603720 | 629207 | 629981 | -0.610 | 11.032 | -3.089 | **0.002** | -0.018 | 0.240 | -0.079 | 0.937 | 0.592 | 5.993 | -2.520 | **0.013** |
| Surface area | 192727 | 198670 | 199488 | -0.489 | 2.214 | -2.473 | **0.015** | -0.059 | 0.243 | -0.260 | 0.795 | 0.430 | 1.361 | -1.827 | 0.070 |
| Intracranial volume | 1507118 | 1578843 | 1574258 | -0.677 | 24.945 | -3.426 | **0.001** | 0.046 | 0.249 | 0.203 | 0.839 | 0.723 | 33.111 | -3.076 | **0.003** |

Follow-up pairwise analysis using t-tests on the estimated marginal means obtained from lsmeans comparing the FHR-SZ vs. PBC, FHR-BP vs. PBC, and FHR-SZ vs. FHR-BP groups in females and males separately. Lsmeans were obtained from ANCOVA models of group differences in brain measures, controlling for age, sex, site, and Euler number. Significant group differences at the uncorrected 0.05 level are shown in bold. Number of male/female children in each group: FHR-SZ: n=51/50, FHR-BP: n=32/32, PBC: n=57/56. Abbreviations: FHR-SZ: Familial high risk for schizophrenia; FHR-BP: Familial high risk for bipolar disorder; PBC: Population-based controls, *d*: Cohen's *d*, $BF_{10}$: Bayes factor.

**Table S8. Results from the pairwise group analysis of the global brain measures in males and females, controlling for axis-I diagnosis.**

| | LS means | | | FHR-SZ vs. PBC | | | FHR-BP vs. PBC | | | FHR-SZ vs. FHR-BP | | |
|---|---|---|---|---|---|---|---|---|---|---|---|---|
| | FHR-SZ | FHR-BP | PBC | *d* | t | p | *d* | t | p | *d* | t | p |
| **Females** | | | | | | | | | | | | |
| Brain volume | 1206868 | 1232973 | 1188631 | 0.207 | 1.014 | 0.569 | 0.504 | 2.256 | 0.066 | -0.297 | -1.274 | 0.413 |
| Cortical volume | 578796 | 594425 | 572156 | 0.154 | 0.754 | 0.732 | 0.517 | 2.313 | 0.058 | -0.363 | -1.557 | 0.268 |
| Surface area | 183079 | 184793 | 179299 | 0.265 | 1.297 | 0.399 | 0.386 | 1.725 | 0.200 | -0.120 | -0.516 | 0.864 |
| Intracranial volume | 1457940 | 1486776 | 1445279 | 0.124 | 0.608 | 0.816 | 0.408 | 1.823 | 0.166 | -0.283 | -1.215 | 0.447 |
| **Males** | | | | | | | | | | | | |
| Brain volume | 1257549 | 1323244 | 1307946 | -0.551 | -2.701 | **0.021** | 0.167 | 0.703 | 0.762 | -0.718 | -3.008 | **0.009** |
| Cortical volume | 604661 | 630531 | 627570 | -0.531 | -2.605 | **0.028** | 0.069 | 0.289 | 0.955 | -0.599 | -2.512 | **0.035** |
| Surface area | 193027 | 198924 | 199303 | -0.450 | -2.210 | 0.073 | -0.027 | -0.114 | 0.993 | -0.423 | -1.774 | 0.183 |
| Intracranial volume | 1509051 | 1580466 | 1570521 | -0.615 | -3.016 | **0.009** | 0.099 | 0.418 | 0.908 | -0.714 | -2.993 | **0.009** |

Follow-up pairwise analysis using t-tests on the estimated marginal means obtained from lsmeans comparing the FHR-SZ vs. PBC, FHR-BP vs. PBC, and FHR-SZ vs. FHR-BP groups in females and males separately. Lsmeans were obtained from ANCOVA models of group differences in brain measures, controlling for age, sex, site, Euler number, and lifetime axis-I diagnosis. Significant group differences at the uncorrected 0.05 level are shown in bold. Number of male/female children in each group: FHR-SZ+: n=23/28, FHR-SZ-: n=27/21, FHR-BP+: n=20/11, FHR-BP-: n=12/21, PBC+: n=16/15, PBC-: n=39/41.Abbreviations: FHR-SZ: Familial high risk for schizophrenia; FHR-BP: Familial high risk for bipolar disorder; PBC: Population-based controls, *d*: Cohen's *d*. Missing data: axis-I disorders PBC males: n=2, FHR-SZ male: n = 1, FHR-SZ female: n=1).

**Table S9. Results from the pairwise analysis comparing participants with and without an axis-I diagnosis in each group.**

| | **LS means** | | | | | | **FHR-SZ+ vs. FHR-SZ-** | | **FHR-BP+ vs. FHR-BP-** | | **PBC+ vs. PBC-** | |
|---|---|---|---|---|---|---|---|---|---|---|---|---|
| | FHR-SZ+ | FHR-SZ- | FHR-BP+ | FHR-BP- | PBC+ | PBC- | t | p | t | p | t | p |
| **Females** | | | | | | | | | | | | |
| Brain volume | 1230796 | 1182862 | 1250021 | 1214056 | 1176199 | 1184049 | 2.029 | **0.049** | 1.006 | 0.324 | -0.287 | 0.775 |
| Cortex volume | 587145 | 569613 | 612152 | 580639 | 564833 | 571513 | 1.491 | 0.143 | 1.811 | 0.081 | -0.516 | 0.608 |
| Surface area | 185634 | 180018 | 187525 | 182492 | 177666 | 179317 | 1.484 | 0.145 | 0.889 | 0.382 | -0.366 | 0.716 |
| Intracranial volume | 1491410 | 1423635 | 1508572 | 1463464 | 1434180 | 1437649 | 2.399 | **0.021** | 1.104 | 0.279 | -0.113 | 0.911 |
| **Males** | | | | | | | | | | | | |
| Brain volume | 1260366 | 1245172 | 1316176 | 1349522 | 1291624 | 1319753 | 0.597 | 0.554 | -0.916 | 0.368 | -0.936 | 0.354 |
| Cortex volume | 605310 | 601180 | 627361 | 643421 | 620614 | 630665 | 0.332 | 0.741 | -0.924 | 0.364 | -0.748 | 0.458 |
| Surface area | 192807 | 192553 | 200113 | 199687 | 198151 | 199645 | 0.059 | 0.953 | 0.087 | 0.931 | -0.332 | 0.741 |
| Intracranial volume | 1508950 | 1499558 | 1569269 | 1604248 | 1568710 | 1574345 | 0.341 | 0.734 | -0.972 | 0.339 | -0.165 | 0.870 |

Follow-up pairwise group analysis comparing the FHR-SZ+ vs. FHR-SZ-, FHR-BP+ vs. FHR-BP-, and PBC+ vs. PBC- groups in females and males separately. Significant group differences at the uncorrected 0.05 level are shown in bold. Number of male/female children in each group: FHR-SZ+: n=23/28, FHR-SZ-: n=27/21, FHR-BP+: n=20/11, FHR-BP-: n=12/21, PBC+: n=16/15, PBC-: n=39/41. Abbreviations: FHR-SZ+: Familial high risk for schizophrenia *with* a lifetime axis-I diagnosis; FHR-SZ-: Familial high risk for schizophrenia *without* a lifetime axis-I diagnosis; FHR-BP+: Familial high risk for bipolar disorder *with* a lifetime axis-I diagnosis; FHR-BP-: Familial high risk for bipolar disorder *without* a lifetime axis-I diagnosis; PBC+: Population-based controls *with* a lifetime axis-I diagnosis; PBC-: Population-based controls *without* a lifetime axis-I diagnosis. Missing data: axis-I disorders PBC males: n=2, FHR-SZ male: n = 1, FHR-SZ female: n=1).
